# Prescription volume and costs of over-the-counter–like drugs in Japan by patient and healthcare facility characteristics: A nationwide descriptive study

**DOI:** 10.64898/2026.01.14.26344144

**Authors:** Yuya Kimura, Akira Okada, Shotaro Aso, Hideo Yasunaga

## Abstract

**Introduction:** “Over-the-counter (OTC)-like drugs” are prescription medicines that contain the same or similar active ingredients as OTC drugs but receive reimbursement under Japan’s universal health insurance system. Despite ongoing policy discussions regarding the potential exclusion of these drugs from insurance coverage or increased patient cost-sharing, evidence describing their real-world prescription volume and costs is limited. We aimed to characterize the use of representative OTC-like drugs in Japan and examine variations by patient and healthcare facility characteristics.

**Methods:** This nationwide descriptive study was conducted using two claims-based data sources. From the National Database of Health Insurance Claims Open Data, we summarized the overall annual prescription volume and costs, further stratified by sex and age group, for fiscal years (FYs) 2018–2023. Using the DeSC database, we performed additional stratified analyses by comorbidity burden, health insurance type, and healthcare facility type. Five OTC-like drug categories were examined: oral single-agent acetaminophen, oral single-agent nonsteroidal anti-inflammatory drugs, topical anti-inflammatory patches, second-generation antihistamines, and heparinoid-containing topical preparations.

**Results:** In FY2023, the five OTC-like drug categories accounted for 17.4 billion prescriptions nationwide (139.6 per capita) and total costs of 288.1 billion yen (2317.0 yen per capita). Heparinoid-containing topical preparations and topical anti-inflammatory patches comprised the largest prescription volumes, whereas second-generation antihistamines and topical anti-inflammatory patches accounted for the highest costs. Prescription volume and costs were higher among females and increased markedly with age, particularly for topical anti-inflammatory patches. Per capita values also increased with greater comorbidity burden, being the highest among individuals covered by the Medical Care System for the Advanced Elderly. Clinics showed higher per capita prescription volume and costs than hospitals.

**Conclusions:** The use of representative OTC-like drugs in Japan is large and varies considerably by patient and healthcare facility characteristics. These findings provide important evidence to inform future policy discussions on OTC-like drugs.

## Introduction

Over-the-counter (OTC) drugs are medications that can be purchased without a prescription, as they have been deemed safe and effective for the self-treatment of mild conditions when used according to label directions^1^. In Japan, there exists a distinct category of medications, called “*OTC-like drugs,*” which generally refer to prescription medications that contain the same or similar active ingredients as OTC drugs but require a physician’s prescription and are reimbursed by public health insurance. These OTC-like drugs include common therapeutic categories such as analgesics, antipyretics, antihistamines, topical anti-inflammatory patches, and moisturizers, many of which are also available without a prescription in other countries.

In Japan, a policy debate has emerged regarding whether OTC-like drugs should be excluded from public health insurance coverage or subject to increased patient copayments^2^. Proponents of this policy argue that excluding these medications from insurance coverage would promote self-medication and reduce national healthcare expenditures, thereby alleviating the financial burden of insurance premiums, particularly for younger populations. However, medical professional organizations and patient advocacy groups have raised concerns about the potential adverse consequences of such policy changes. These include increased out-of-pocket costs for patients who would need to purchase more expensive OTC alternatives, reduced access to necessary medications, particularly among economically or medically vulnerable individuals, and a potential increase in disease progression resulting from delayed or forgone healthcare consultations. Despite the ongoing policy discussion, evidence regarding the prescription volume and costs of OTC-like drugs in Japan is limited, including how these prescriptions are distributed across patient subgroups with varying comorbidity burdens and across different types of healthcare facilities. Such evidence is essential for informing evidence-based policy decisions and understanding the potential implications of insurance coverage changes on specific patient populations and healthcare settings.

To address this evidence gap, we aimed to describe the prescription volume and costs of representative OTC-like drugs in Japan, both overall and stratified by patient and healthcare facility characteristics. Specifically, our objectives were as follows: 1) to quantify the prescription volume and costs for each drug category; 2) to stratify these measures by patient characteristics, including sex, age group, Charlson Comorbidity Index (CCI) category^3^, the presence of comorbidities of major causes of death in Japan, and health insurance type; and 3) to examine the prescription volume and costs across healthcare facility types, including clinics, non-university hospitals, and university hospitals.

## Methods

### Data source

Two data sources were used for this study: 1) the National Database of Health Insurance Claims (NDB) Open Data and 2) the DeSC database. The NDB is a nationwide administrative claims database covering approximately 99% of hospitals in Japan and contains various types of information, including age, sex, disease diagnoses coded according to the International Classification of Diseases, 10^th^ Revision (ICD-10), procedures, and medications in both inpatient and outpatient settings^4^. Although the NDB is an important data source for epidemiological research, access is limited due to a multistep application process that requires a certain amount of time^5^. In parallel, the Ministry of Health, Labour, and Welfare of Japan, which administers the NDB, provides an open-access, publicly available version known as the NDB Open Data^4^. While the NDB Open Data does not include patient- or facility-level information, it contains aggregated medication data, including prescription volumes for each fiscal year (FY), for both the overall database population and stratified by sex and age group.

The DeSC database is a large-scale health insurance claims database in Japan^5,6^. Five types of public health insurance are operational in Japan: 1) the municipality-based National Health Insurance for non-employees; 2) Japan Health Insurance Association-managed insurance for employees of small companies; 3) association- or union-managed insurance for employees of large companies; 4) Mutual Aid Association-managed insurance for civil servants; and 5) the Advanced Elderly Medical Service System for individuals aged 75 years or above. While the NDB consolidates information from all five insurance types, the DeSC database collects information from the first, third, and fifth types. The data elements contained in the DeSC database are similar to those in the NDB.

We selected the NDB Open Data as the primary data source to quantify national prescription volumes and costs for each study drug and stratify these data by sex and age group. Because the NDB Open Data provides aggregated data without information required for more detailed stratification, prescription volumes and costs could not be further stratified by CCI category, the presence of comorbid conditions corresponding to major causes of death in Japan, insurance type, or healthcare facility type. We used the DeSC database to enable these stratified analyses. For both the NDB Open Data and DeSC database, we extracted data from FYs 2018 through 2023 (April 2018 to March 2024).

### Study drugs

We focused on five representative OTC-like drug categories: oral single-agent acetaminophen, oral single-agent nonsteroidal anti-inflammatory drugs (NSAIDs), topical anti-inflammatory patches, oral second-generation antihistamines, and heparinoid-containing topical preparations. The analysis was limited to outpatient prescriptions because inpatient drug costs are often integrated into bundled reimbursement systems, making it difficult to isolate and calculate the specific costs of individual drugs. The WHO-ATC codes for each drug category are provided in Supplementary Table 1.

### Analysis using the NDB Open Data

Using information extracted from the NDB Open Data, we calculated the annual total prescription volume and costs for each study drug category for each FY. We further stratified these data by sex and age group (≤64, 65–74, and ≥75 years). Additionally, we calculated the per capita prescription volume and costs for each study drug. For these calculations, the total prescription volume and costs for each study drug category in each FY were divided by either the total estimated population of Japan or the estimated number of individuals in each stratum in that year^7^.

### Analysis using the DeSC database

Using the DeSC database, we first calculated the total prescription volume and costs for each study drug category in each FY, to describe the overall prescription patterns captured in the DeSC database and facilitate comparison with the NDB Open Data. Thereafter, we stratified these data by CCI category (0, 1–2, and ≥3), presence of one of the leading causes of death in Japan, health insurance type, and healthcare facility type.

The CCI is a weighted score that sums assigned points for 17 categories of chronic conditions and can predict both short-term and long-term outcomes, including mortality and functional status^3,8^. The ICD-10 codes for the 17 chronic conditions were obtained from a previous study^8^. We limited our analysis to chronic conditions, viz. malignant neoplasms, heart diseases, cerebrovascular diseases, renal diseases, and Alzheimer’s disease, from among Japan’s ten leading causes of death in 2024^9^. The ICD-10 codes for these conditions were defined in accordance with the national mortality survey and are presented in Supplementary Table 2^10^. A condition was considered to be present if the corresponding diagnostic code appeared during the FY, both for calculating the CCI and identifying chronic conditions. Health insurance types included the National Health Insurance, Health Insurance for employees of large companies, and the Medical Care System for the Advanced Elderly. Healthcare facilities were categorized into clinics, non-university hospitals, and university hospitals.

For the stratified analyses, we also calculated the per capita prescription volume and costs for each study drug. For these calculations, the total prescription volume and costs for each study drug category in each FY were divided by the corresponding denominator population in the year: individuals with a CCI of 0, 1–2, or ≥3; individuals with each of the above-mentioned chronic conditions; individuals enrolled in each insurance type; and individuals who visited each type of facility. A separate denominator was estimated for the respective FY. In the stratified analyses, results were primarily reported as per capita values for strata with substantial differences in population size, to ensure interpretable comparisons.

To evaluate the representativeness of the individual composition within the DeSC database relative to the general Japanese population, we summarized the number and proportion of individuals by health insurance type in the DeSC database. According to national statistics reported in Japan in 2022, the nationwide distribution was as follows: National Health Insurance (25.4 million; 20.9%), Health Insurance for employees of small companies (40.3 million; 33.2%), Health Insurance for employees of large companies (28.4 million; 23.4%), Mutual Aid Associations for civil servants (8.7 million; 7.2%), and Medical Care System for the Advanced Elderly (18.4 million; 15.2%)^11^.

## Results

### Results from the NDB Open Data

#### Overall prescription volume and costs

The ranking of OTC-like drug categories by total prescription volume changed over time (Table 1 and Figure 1). In FY2018, the three most frequently prescribed drug categories were topical anti-inflammatory patches, heparinoid-containing topical preparations, and second-generation antihistamines. By FY2023, this ranking shifted to heparinoid-containing topical preparations, topical anti-inflammatory patches, and second-generation antihistamines. In FY2023, the total prescription volumes (per capita values) of the three leading categories were 5.8 billion (46.9), 4.4 billion (35.5), and 3.9 billion (31.7) for heparinoid-containing topical preparations, topical anti-inflammatory patches, and second-generation antihistamines, respectively. The total volume (per capita value) for the five study drug categories in FY2023 was 17.4 billion (139.6).

**Figure 1.**
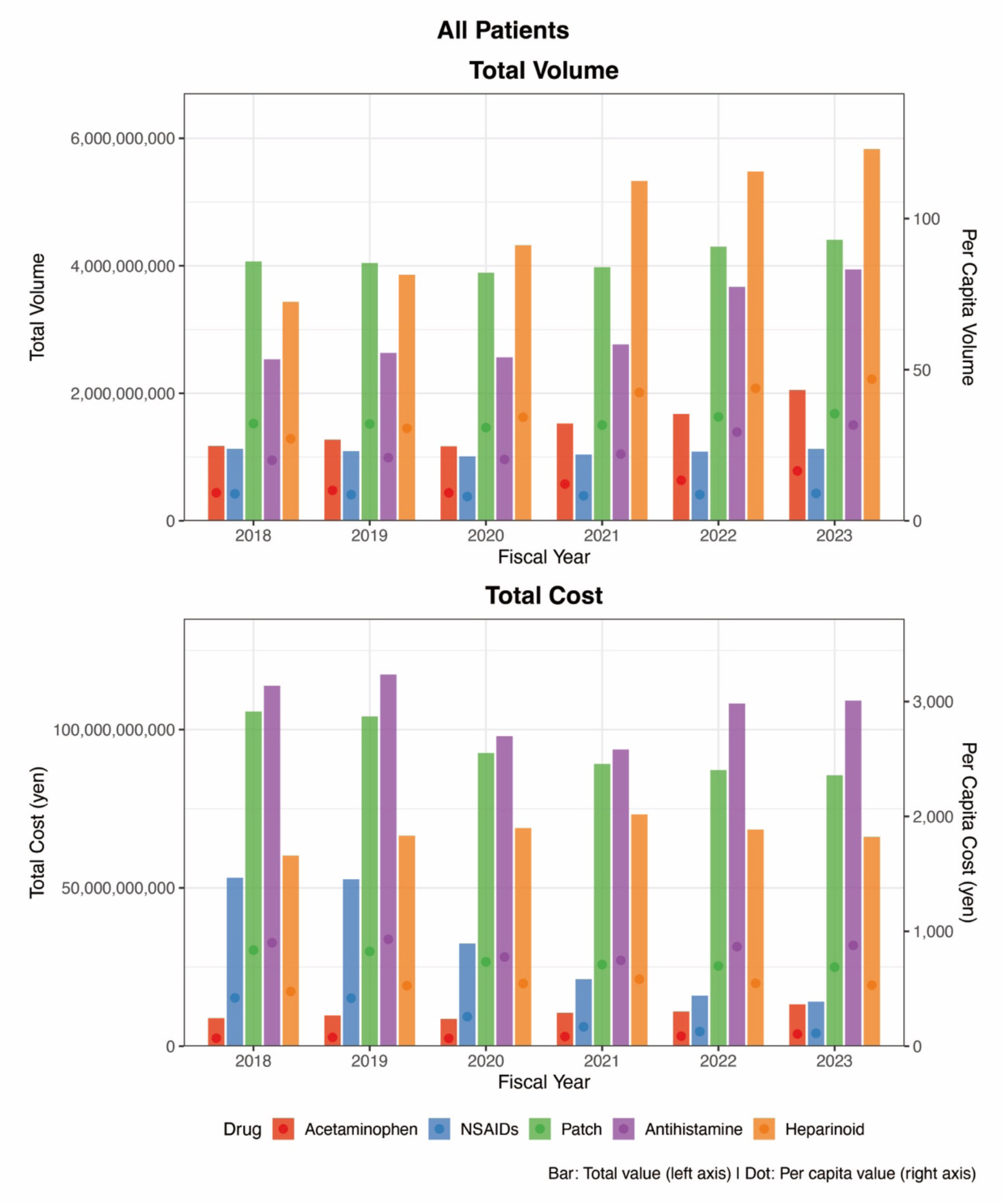
Trends in prescription volume and costs of representative OTC-like drugs in the overall population: Analysis of the NDB Open Data. The upper panel depicts the total prescription volume (bars, left y-axis) and per capita prescription volume (dots, right y-axis), and the lower panel depicts the total prescription costs (bars, left y-axis) and per capita prescription costs (dots, right y-axis). Note: Prescription volume and costs are presented for oral single-agent acetaminophen, oral single-agent nonsteroidal anti-inflammatory drugs (NSAIDs), topical anti-inflammatory patches, oral second-generation antihistamines, and heparinoid-containing topical preparations. The per capita values were calculated by dividing the annual totals by the estimated Japanese population in each fiscal year.

**Table 1.**
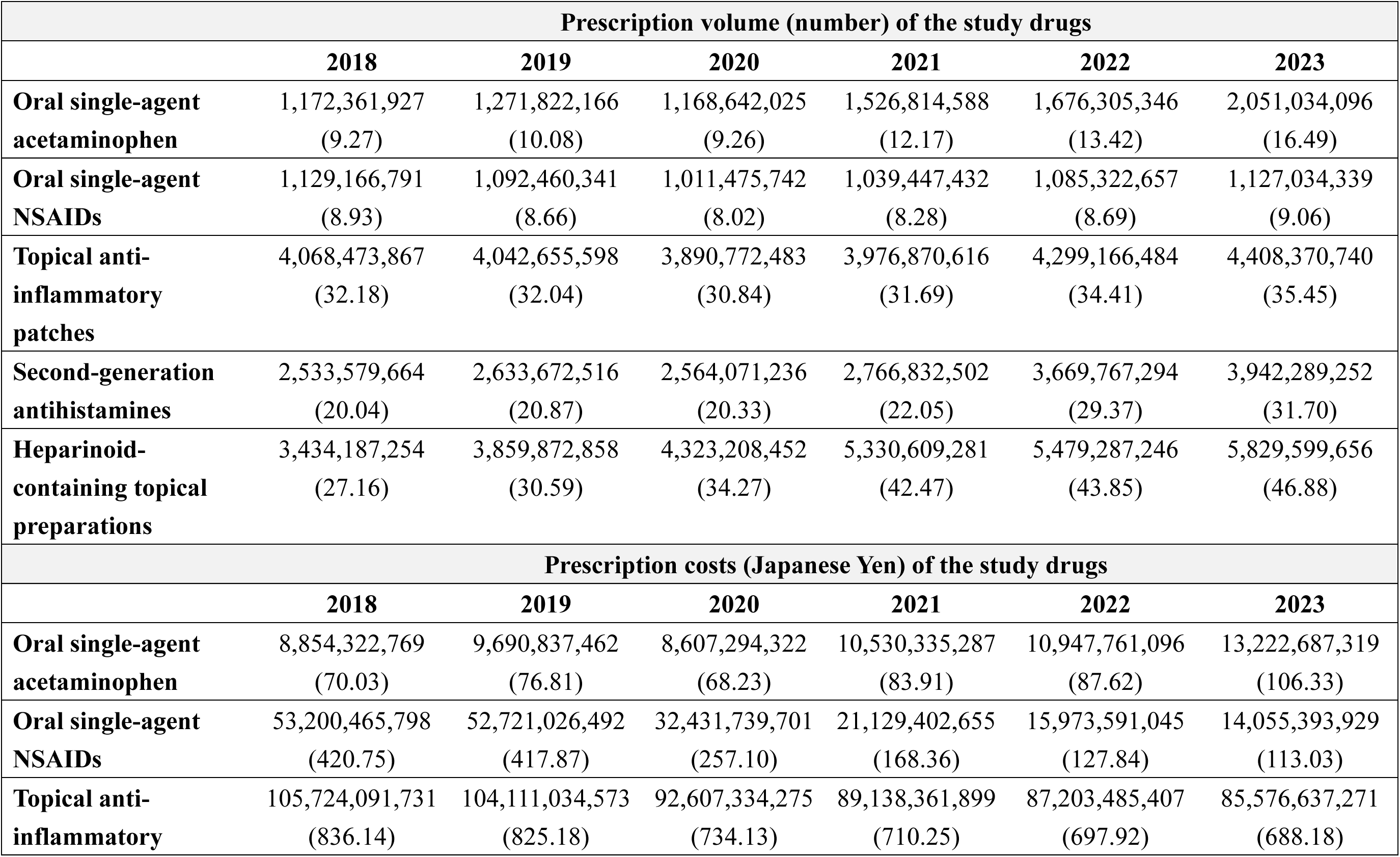
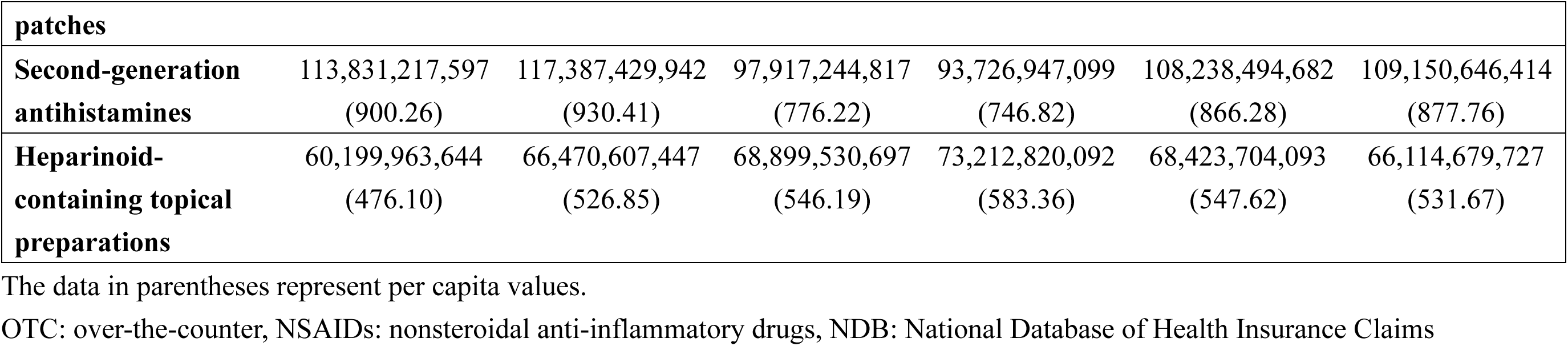
Annual prescription volume and costs for representative OTC-like drugs: Analysis of the NDB Open Data.

In contrast, the ranking of drug categories by total prescription costs remained unchanged throughout the study period, with second-generation antihistamines, topical anti-inflammatory patches, and heparinoid-containing topical preparations consistently accounting for the highest costs. In FY2023, the total costs (per capita values) were 109.2 billion yen (877.8 yen) for second-generation antihistamines, 85.6 billion yen (688.2 yen) for topical anti-inflammatory patches, and 66.1 billion yen (531.7 yen) for heparinoid-containing topical preparations. The total costs (per capita values) for the five study drug categories in FY2023 were 288.1 billion yen (2317.0 yen).

#### Sex-specific prescription volume and costs

Across all fiscal years and all drug categories, females consistently had higher total prescription volume and total costs than males, both in absolute terms and on a per capita basis (Supplementary Table 3, Figure 2).

**Figure 2.**
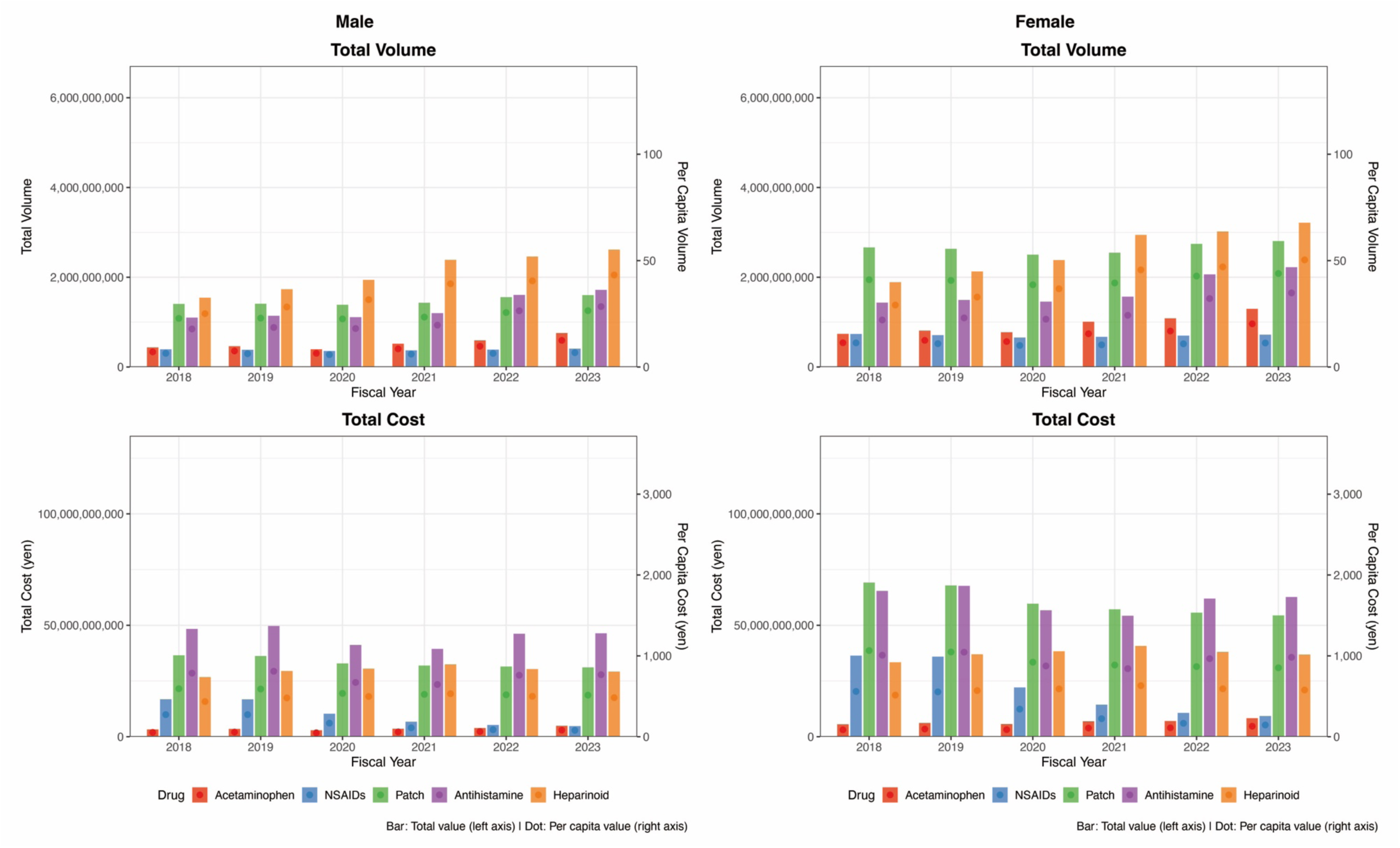
Sex-specific trends in prescription volume and costs of representative OTC-like drugs: Analysis of the NDB Open Data. (A) Trends in the total prescription volume (bars, left y-axis) and per capita prescription volume (dots, right y-axis) among males. (B) Trends in the total prescription volume (bars, left y-axis) and per capita prescription volume (dots, right y-axis) among. Note: Prescription volume and costs are presented for oral single-agent acetaminophen, oral single-agent nonsteroidal anti-inflammatory drugs (NSAIDs), topical anti-inflammatory patches, oral second-generation antihistamines, and heparinoid-containing topical preparations.

In FY2023, the three leading drug categories by total prescription volume differed by sex. Among males, the top three categories were heparinoid-containing topical preparations, second-generation antihistamines, and topical anti-inflammatory patches, with total volumes (per capita values) of 2.6 billion (43.2), 1.7 billion (28.4), and 1.6 billion (26.5), respectively. In contrast, among females, the top three categories were heparinoid-containing topical preparations, topical anti-inflammatory patches, and second-generation antihistamines, with corresponding values of 3.2 billion (50.3), 2.8 billion (43.9), and 2.2 billion (34.8). The total volume (per capita value) for the five study drug categories in FY2023 was 7.1 billion (117.3) for males, and 10.3 billion (160.6) for females.

The top three drug categories accounting for the majority of the total prescription costs in FY2023 were identical for males and females: second-generation antihistamines, topical anti-inflammatory patches, and heparinoid-containing topical preparations. Among males, the total costs (per capita values) were 46.4 billion yen (767.5 yen) for antihistamines, 31.1 billion yen (514.3 yen) for patches, and 29.2 billion yen (483.4 yen) for heparinoids. The corresponding values among females were 62.7 billion yen (981.7 yen), 54.5 billion yen (852.7 yen), and 36.9 billion yen (577.5 yen), respectively. The total costs (per capita values) for the five study drug categories in FY2023 were 116.5 billion yen (1925.3 yen) for males, and 171.6 billion yen (2687.3 yen) for females.

#### Age-group-specific prescription volume and costs

In FY2023, the drug category with the highest per capita prescription volume varied by age group (Supplementary Table 4, Figure 3). Among individuals aged 0–64 years, the per capita volume (48.6) was the highest for heparinoid-containing topical preparations. Among those aged 65–74 years and ≥75 years, topical anti-inflammatory patches ranked the highest, with per capita volumes of 61.7 and 118.9, respectively. The total per capita volume for the five study drug categories in FY2023 were 106.7 in the 0–64 years age group, 154.7 in the 65–74 years group, and 271.8 in the ≥75 years group.

**Figure 3.**
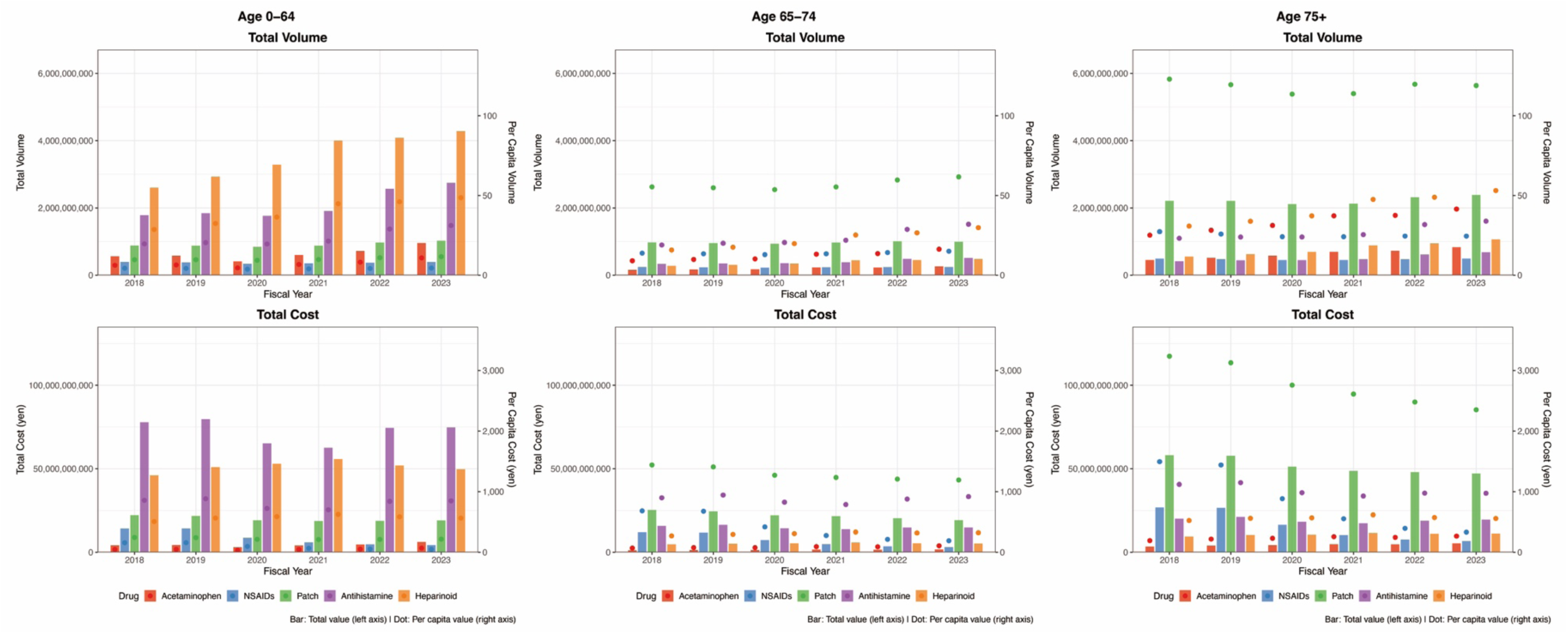
Age-group–specific trends in prescription volume and costs of representative OTC-like drugs: Analysis of the NDB Open Data. (A) Trends in total prescription volume (bars, left y-axis) and per capita prescription volume (dots, right y-axis) among individuals aged 0–64 years. (B) Trends in total prescription volume (bars, left y-axis) and per capita prescription volume (dots, right y-axis) among individuals aged 65–74 years. (C) Trends in total prescription volume (bars, left y-axis) and per capita prescription volume (dots, right y-axis) among individuals aged ≥75 years. Note: Prescription volume and costs are presented for oral single-agent acetaminophen, oral single-agent nonsteroidal anti-inflammatory drugs (NSAIDs), topical anti-inflammatory patches, oral second-generation antihistamines, and heparinoid-containing topical preparations.

The leading drug category by per capita prescription costs also differed by age group. In the 0–64 years age group, second-generation antihistamines accounted for the highest per capita cost (848.1 yen), whereas topical anti-inflammatory patches accounted for the highest per capita costs in the 65–74 year and ≥75 year age groups (1192.6 yen and 2351.2 yen, respectively). The total costs per capita for the five study drug categories in FY2023 were 1747.8 yen in the 0–64 years age group, 2729.4 yen in the 65–74 years group, and 4481.2 yen in the ≥75 years group.

Comparison of the youngest (0–64 years) and oldest (≥75 years) age groups revealed that the per capita prescription volume and costs for topical anti-inflammatory patches were more than tenfold higher in the oldest group, while the corresponding per capita values for second-generation antihistamines and heparinoid-containing topical preparations were similar for these age groups.

### Results from the DeSC database

#### Overall prescription volume and costs

In the analysis of the whole population from the DeSC database, the ranking of OTC-like drug categories by total prescription volume and total costs differed from that observed in the NDB Open Data (Supplementary Table 5 and Supplementary Figure 1). Additionally, the per capita prescription volumes and costs were consistently smaller than those estimated from the NDB Open Data.

In FY2023, the three leading drug categories by total prescription volume were topical anti-inflammatory patches, heparinoid-containing topical preparations, and second-generation antihistamines, with total volumes (per capita values) of 80.0 million (8.5), 25.0 million (2.7), and 23.1 million (2.5), respectively. In contrast, the ranking by total prescription costs was as follows: topical anti-inflammatory patches, second-generation antihistamines, and heparinoid-containing topical preparations, with corresponding costs (per capita values) of 1.6 billion yen (170.6 yen), 0.68 billion yen (72.2 yen), and 0.33 billion yen (35.1 yen).

The total volume (per capita value) and costs (per capita value) for the five study drug categories in FY2023 were 155.3 million (16.5) and 2.94 billion yen (312.1 yen), respectively.

#### CCI-specific prescription volume and costs

Across all study drug categories, both per capita total prescription volume and costs increased with higher CCI categories (Supplementary Table 6 and Supplementary Figure 2).

The total volume per capita for the five study drug categories in FY2023 were 6.9 in the CCI=0 category, 36.7 in the CCI=1–2 category, and 49.1 in the CCI ≥3 category. Moreover, the total per capita costs for the five study drug categories in FY2023 were 130.7 yen in the CCI=0 category, 699.3 yen in the CCI=1–2 category, and 915.1 in the CCI ≥3 category.

#### Major-disease–specific prescription volume and costs

The total per capita volume for the five study drug categories in FY2023 was 45.2 in patients with malignant neoplasms, 47.8 in patients with heart diseases, 43.0 in patients with cerebrovascular diseases, 47.9 in patients with renal diseases, and 27.4 in patients with Alzheimer’s disease. Additionally, the total per capita costs for the five study drug categories in FY2023 were 837.4 yen for patients with malignant neoplasms, 898.4 yen for patients with heart diseases, 808.1 yen for patients with cerebrovascular diseases, 879.7 yen for patients with renal diseases, and 493.3 yen for patients with Alzheimer’s disease (Supplementary Table 7 and Supplementary Figure 3).

#### Health-insurance-specific prescription volume and costs

National Health Insurance accounted for approximately 45–55%, Health Insurance for approximately 7–10%, and the Medical Care System for the Advanced Elderly for approximately 35–45% of individuals in the DeSC database (Supplementary Table 8).

The total volume per capita for the five study drug categories in FY2023 was 7.0 in individuals covered by the National Health Insurance, 12.3 in individuals covered by Health Insurance, and 26.4 in individuals covered by the Medical Care System for the Advanced Elderly (Supplementary Table 9 and Supplementary Figure 4). Additionally, the total costs per capita for the five study drug categories in FY2023 were 131.8 yen for individuals covered by the National Health Insurance, 227.5 yen for individuals covered by Health Insurance, and 499.2 yen for individuals covered by the Medical Care System for the Advanced Elderly.

#### Healthcare-facility–specific prescription volume and costs

The total per capita volume for the five study drug categories in FY2023 was 30.9, 11.6, and 12.1 for individuals visiting clinics, non-university hospitals, and university hospitals, respectively (Supplementary Table 9 and Supplementary Figure 5). The total costs per capita for the five study drug categories in FY2023 were 589.3, 209.9, and 232.8 yen for individuals visiting clinics, non-university hospitals, and university hospitals, respectively.

## Discussion

Using both nationwide claims data in an aggregated manner and individual-level data, this study provides a contemporary comprehensive description of the scale and distribution of prescription use of representative OTC-like drugs in Japan. We found that these drugs accounted for a substantial volume of prescriptions and healthcare expenditures, with an annual national prescription volume of approximately 17.4 billion prescriptions and total costs of 288.1 billion yen. Among the five drug categories examined, heparinoid-containing topical preparations, topical anti-inflammatory patches, and second-generation antihistamines accounted for the largest prescription volumes, whereas second-generation antihistamines and topical anti-inflammatory patches contributed the most to the total costs.

The prescription volume and cost varied markedly across population subgroups. Females consistently had higher per capita prescription volume and costs than males, and both measures increased substantially with advancing age. In particular, the per capita prescription volume and costs for topical anti-inflammatory patches were more than ten times higher among individuals aged ≥75 years compared with those aged 0–64 years, whereas age-related differences were less pronounced for second-generation antihistamines and heparinoid-containing topical preparations.

Several features of the observed prescription patterns warrant consideration. The higher prescription volume and costs among older adults and individuals with a greater comorbidity burden likely reflect the higher prevalence of chronic symptoms and conditions for which these OTC-like drugs are commonly prescribed, such as pain, inflammation, allergic symptoms, and skin disorders. The marked age-related increase in the use of topical anti-inflammatory patches may be attributable to the high burden of musculoskeletal conditions in older populations^12^ as well as the clinical preference for topical agents to reduce the risk of systemic adverse effects. This preference may be reflected in the substantially higher prescription volume of topical anti-inflammatory patches compared with oral NSAIDs among older adults. In contrast, the relatively modest age-related differences for second-generation antihistamines and heparinoid-containing topical preparations may reflect their use across a broader age range for conditions that are less strongly age-dependent.

Differences by healthcare facility type may also reflect variations in clinical practice patterns and patient case mix. The higher prescription volume and costs observed in clinics, compared with hospitals, may be related to the role of clinics as primary points of contact for patients with chronic, non-life-threatening conditions, for which OTC-like drugs are frequently prescribed. Similarly, differences across health insurance types are likely influenced by underlying differences in age distribution and comorbidity burden among insured populations, rather than by the type of insurance itself.

Amid the ongoing discussion about increasing patient cost-sharing for selected OTC-like drugs, the description of national prescription volume and costs across population subgroups provided by this study may serve as a useful reference for policy considerations. If patient cost-sharing were increased, the financial impact would not be uniform across the population, given the substantial differences in prescription volume and costs observed by age, comorbidity burden, and insurance type. Conversely, if current coverage policies remain unchanged, the observed trends may persist, with continued implications for healthcare expenditure. Importantly, the present study did not evaluate the effects of cost-sharing policies on prescribing behavior or health outcomes. If policy changes regarding patient cost-sharing for OTC-like drugs are implemented in the future, the present findings may serve as baseline data for evaluating subsequent changes in prescription patterns and healthcare expenditures.

This study has some limitations. First, some prescription volume values—generally those with fewer than 1000 prescriptions—were masked in the NDB Open Data, leading to slight underestimation of the prescription volume and costs. However, because these masked values accounted for only a very small proportion of total prescriptions, the magnitude of underestimation is likely minimal. Second, individuals included in the DeSC database may not be representative of the overall Japanese population. The proportion of individuals covered by the Medical Care System for the Advanced Elderly was higher in the DeSC database than in national statistics (35–45% vs. 15.2%), resulting in overrepresentation of older adults. This likely explains why the overall prescription volume and cost distributions observed in the DeSC database (Supplementary Figure 1) resembled those among individuals aged ≥75 years in the NDB Open Data (Figure 3). Nevertheless, despite its limited representativeness for national-level estimates, the analyses of DeSC database provide valuable insights into differences in prescription patterns across patient subgroups and healthcare settings.

In conclusion, using the NDB Open Data, we showed that the five representative OTC-like drug categories accounted for 288.1 billion yen (2317.0 yen per capita) in prescription costs in fiscal year 2023. The three drug categories that contributed to the largest share of the total costs were second-generation antihistamines (109.2 billion yen; 877.8 yen per capita), topical anti-inflammatory patches (85.6 billion yen; 688.2 yen per capita), and heparinoid-containing topical preparations (66.1 billion yen; 531.7 yen per capita). The magnitude and composition of OTC-like drug use varied substantially by sex and age group.

## Data Availability

NDB Open Data is publicly accessible. In addition, the DeSC database is available for commercial purchase.

https://www.mhlw.go.jp/stf/seisakunitsuite/bunya/0000177182.html

## Acknowledgements

The authors would like to thank Mr. Masayoshi Kurihara, a Health Information Manager at National Hospital Organization Tokyo National Hospital, for his helpful advice and information regarding OTC-like drugs. The authors used ChatGPT 5.2 (by OpenAI) and Gemini 3 (by Google) to enhance the readability of the English text during the preparation of this article. After using these tools, the authors reviewed and edited the content as needed and take full responsibility for the publication.

## Author contributions

YK designed the research, collected the data, performed statistical analysis, and wrote the first draft. AO and AS designed the research and revised the first draft. HY revised the first draft and supervised the entire manuscript preparation. All authors have read and approved the final manuscript.

## Approval by institutional review board

This study protocol was approved by the Institutional Review Board of the Graduate School of Medicine of the University of Tokyo [approval number: 2021010NI (April 23, 2021)], and the study was conducted in accordance with the principles of the Helsinki Declaration.

## List of Supplementary Materials

**Supplementary Table 1.** WHO–ATC codes for the study drugs.

| Study drugs | WHO-ATC codes |
| --- | --- |
| Oral single-agent acetaminophen | N02BE01 |
| Oral single-agent NSAIDs | M01AB, M01AC, M01AE, M01AG, M01AH |
| Topical anti-inflammatory patches* | M02AA, M02AB, M02AC, M02AX |
| Second-generation antihistamines** | R06AX24, R06AX22, R06AE07, R06AX26, R01AC08, R06AX13, R06AE09, R06AX29, R06AX27, R06AX28 |
| Heparinoid-containing topical preparations*** | Not applicable |

NSAIDs, nonsteroidal anti-inflammatory drugs; WHO, World Health Organization; ATC, Anatomical Therapeutic Chemical classification
\*Because some topical agents with the listed WHO-ATC codes are not patches, we have included only the products specifically categorized as patches.
\*\*Bepotastine is a second-generation antihistamine used in Japan; however, no WHO-ATC code is assigned to this drug. Therefore, bepotastine was included in the second-generation antihistamines category based on its drug name rather than the WHO–ATC code.
\*\*\*No WHO–ATC codes are assigned to heparinoid-containing topical preparations. Therefore, we included in this category those drugs whose product name contains “hirudoid” or “heparinoid.”

**Supplementary Table 2.** ICD-10 codes for the leading causes of death.

| Disease category | ICD-10 codes |
| --- | --- |
| Malignant neoplasms | C00–97 |
| Heart diseases | I01–020, I05–09, I20–25, I27, I30–52 |
| Cerebrovascular diseases | I60–69 |
| Renal diseases | N17–19 |
| Alzheimer disease | G30 |

ICD-10, International Classification of Diseases, 10<sup>th</sup> Revision

**Supplementary Table 3.** Annual prescription volume and costs of representative OTC-like drugs stratified by sex: Analysis of the NDB Open Data.

| **Male** |  |  |  |  |  |  |
| --- | --- | --- | --- | --- | --- | --- |
| **Prescription volume (number) of the study drugs** |  |  |  |  |  |  |
|  | **2018** | **2019** | **2020** | **2021** | **2022** | **2023** |
| **Oral single-agent acetaminophen** | 434,302,072<br>(7.06) | 462,284,555<br>(7.53) | 394,241,983<br>(6.43) | 519,119,445<br>(8.51) | 592,389,856<br>(9.75) | 756,268,701<br>(12.50) |
| **Oral single-agent NSAIDs** | 393,861,956<br>(6.40) | 382,550,633<br>(6.23) | 356,064,746<br>(5.80) | 368,317,556<br>(6.04) | 387,248,060<br>(6.37) | 407,345,728<br>(6.73) |
| **Topical anti-inflammatory patches** | 1,405,109,216<br>(22.84) | 1,409,567,077<br>(22.95) | 1,386,794,264<br>(22.60) | 1,430,406,678<br>(23.44) | 1,554,860,554<br>(25.59) | 1,601,518,170<br>(26.47) |
| **Second-generation antihistamines** | 1,099,829,456<br>(17.87) | 1,140,519,149<br>(18.57) | 1,108,920,552<br>(18.08) | 1,199,696,921<br>(19.66) | 1,605,496,489<br>(26.42) | 1,717,876,172<br>(28.40) |
| **Heparinoid-containing topical preparations** | 1,543,834,983<br>(25.09) | 1,732,173,388<br>(28.21) | 1,941,248,831<br>(31.64) | 2,387,082,394<br>(39.12) | 2,459,930,507<br>(40.49) | 2,614,579,277<br>(43.22) |
| **Prescription costs (Japanese Yen) of the study drugs** |  |  |  |  |  |  |
|  | **2018** | **2019** | **2020** | **2021** | **2022** | **2023** |
| **Oral single-agent acetaminophen** | 3,277,761,809<br>(53.27) | 3,523,113,513<br>(57.37) | 2,918,084,956<br>(47.56) | 3,601,031,329<br>(59.01) | 3,888,723,051<br>(64.00) | 4,912,998,563<br>(81.22) |
| **Oral single-agent NSAIDs** | 16,820,841,878<br>(273.37) | 16,783,061,930<br>(273.29) | 10,313,644,080<br>(168.11) | 6,736,943,463<br>(110.41) | 5,288,275,611<br>(87.04) | 4,773,158,982<br>(78.91) |
| **Topical anti-inflammatory patches** | 36,534,662,734<br>(593.75) | 36,233,045,296<br>(590.01) | 32,890,288,606<br>(536.11) | 31,946,410,369<br>(523.55) | 31,517,633,932<br>(518.74) | 31,110,749,791<br>(514.30) |
| **Second-generation antihistamines** | 48,384,127,421<br>(786.32) | 49,688,790,377<br>(809.12) | 41,184,273,165<br>(671.30) | 39,426,021,127<br>(646.13) | 46,225,133,211<br>(760.81) | 46,427,641,534<br>(767.50) |
| **Heparinoid-containing topical preparations** | 26,795,649,736<br>(435.48) | 29,496,514,954<br>(480.31) | 30,555,068,767<br>(498.05) | 32,466,105,662<br>(532.07) | 30,364,039,739<br>(499.75) | 29,238,721,284<br>(483.35) |
| **Female** |  |  |  |  |  |  |
| **Prescription volume (number) of the study drugs** |  |  |  |  |  |  |
|  | **2018** | **2019** | **2020** | **2021** | **2022** | **2023** |
| **Oral single-agent acetaminophen** | 738,034,717<br>(11.37) | 809,494,871<br>(12.50) | 774,379,776<br>(11.95) | 1,007,663,087<br>(15.63) | 1,083,883,388<br>(16.89) | 1,294,743,841<br>(20.28) |
| **Oral single-agent NSAIDs** | 735,209,968<br>(11.33) | 709,845,879<br>(10.96) | 655,335,211<br>(10.11) | 671,045,989<br>(10.41) | 697,977,069<br>(10.87) | 719,599,875<br>(11.27) |
| **Topical anti-inflammatory patches** | 2,663,290,851<br>(41.03) | 2,633,020,569<br>(40.66) | 2,503,911,937<br>(38.64) | 2,546,403,101<br>(39.49) | 2,743,450,126<br>(42.74) | 2,806,000,301<br>(43.94) |
| **Second-generation antihistamines** | 1,433,690,610<br>(22.09) | 1,493,106,014<br>(23.06) | 1,455,068,459<br>(22.46) | 1,567,044,165<br>(24.30) | 2,062,846,425<br>(32.14) | 2,222,900,380<br>(34.81) |
| **Heparinoid-containing topical preparations** | 1,890,263,091<br>(29.12) | 2,127,616,138<br>(32.86) | 2,381,814,241<br>(36.76) | 2,943,413,824<br>(45.65) | 3,019,234,285<br>(47.04) | 3,214,887,643<br>(50.34) |
| **Prescription costs (Japanese Yen) of the study drugs** |  |  |  |  |  |  |
|  | **2018** | **2019** | **2020** | **2021** | **2022** | **2023** |
| **Oral single-agent acetaminophen** | 5,576,360,944<br>(85.91) | 6,167,427,130<br>(95.24) | 5,689,064,224<br>(87.80) | 6,929,106,981<br>(107.46) | 7,058,836,105<br>(109.97) | 8,309,544,849<br>(130.12) |
| **Oral single-agent NSAIDs** | 36,378,066,529<br>(560.43) | 35,936,823,623<br>(554.96) | 22,116,691,173<br>(341.32) | 14,391,264,469<br>(223.18) | 10,684,148,845<br>(166.45) | 9,281,367,986<br>(145.34) |
| **Topical anti-inflammatory patches** | 69,188,223,487<br>(1065.89) | 67,876,731,595<br>(1048.19) | 59,715,802,968<br>(921.58) | 57,190,852,187<br>(886.91) | 55,673,030,692<br>(867.33) | 54,453,091,493<br>(852.71) |
| **Second-generation antihistamines** | 65,444,274,687<br>(1008.22) | 67,696,650,341<br>(1045.41) | 56,730,668,183<br>(875.51) | 54,297,955,963<br>(842.05) | 61,979,793,034<br>(965.58) | 62,690,440,534<br>(981.70) |
| **Heparinoid-containing topical preparations** | 33,403,590,077<br>(514.61) | 36,973,402,331<br>(570.96) | 38,343,188,928<br>(591.74) | 40,745,869,942<br>(631.89) | 38,058,714,764<br>(592.92) | 36,875,126,583<br>(577.45) |

The data in parentheses represent per capita values.
OTC: over-the-counter, NDB: National Database of Health Insurance Claims, NSAIDs: nonsteroidal anti-inflammatory drugs

**Supplementary Table 4.** Annual prescription volume and costs of representative OTC-like drugs stratified by age: Analysis of the NDB Open Data.

| **Age ≤64 years** |  |  |  |  |  |  |
| --- | --- | --- | --- | --- | --- | --- |
| **Prescription volume (number) of the study drugs** |  |  |  |  |  |  |
|  | **2018** | **2019** | **2020** | **2021** | **2022** | **2023** |
| **Oral single-agent acetaminophen** | 562,515,986<br>(6.19) | 579,705,445<br>(6.42) | 410,573,984<br>(4.56) | 601,194,602<br>(6.73) | 721,787,643<br>(8.14) | 955,506,716<br>(10.84) |
| **Oral single-agent NSAIDs** | 394,844,792<br>(4.35) | 382,191,871<br>(4.23) | 338,421,258<br>(3.76) | 353,668,303<br>(3.96) | 370,861,732<br>(4.18) | 392,743,389<br>(4.46) |
| **Topical anti-inflammatory patches** | 882,676,071<br>(9.71) | 879,161,485<br>(9.74) | 843,386,926<br>(9.36) | 879,309,821<br>(9.85) | 971,442,433<br>(10.95) | 1,024,033,177<br>(11.62) |
| **Second-generation antihistamines** | 1,784,923,105<br>(19.64) | 1,843,085,929<br>(20.42) | 1,761,328,192<br>(19.54) | 1,907,730,413<br>(21.37) | 2,569,311,886<br>(28.96) | 2,745,318,179<br>(31.15) |
| **Heparinoid-containing topical preparations** | 2,603,314,010<br>(28.65) | 2,929,772,573<br>(32.45) | 3,286,576,418<br>(36.47) | 4,001,216,968<br>(44.81) | 4,085,121,555<br>(46.05) | 4,282,891,723<br>(48.60) |
| **Prescription costs (Japanese Yen) of the study drugs** |  |  |  |  |  |  |
|  | **2018** | **2019** | **2020** | **2021** | **2022** | **2023** |
| **Oral single-agent acetaminophen** | 4,182,482,132<br>(46.03) | 4,343,349,046<br>(48.11) | 2,993,967,873<br>(33.22) | 4,106,926,352<br>(46.00) | 4,668,225,666<br>(52.62) | 6,158,537,509<br>(69.88) |
| **Oral single-agent NSAIDs** | 14,293,941,753<br>(157.31) | 14,314,684,255<br>(158.56) | 8,693,439,106<br>(96.47) | 5,953,119,481<br>(66.67) | 4,740,106,414<br>(53.43) | 4,320,269,209<br>(49.02) |
| **Topical anti-inflammatory patches** | 22,218,337,424<br>(244.51) | 21,793,753,385<br>(241.40) | 19,161,795,980<br>(212.63) | 18,739,005,100<br>(209.87) | 18,806,373,469<br>(212.00) | 19,104,112,886<br>(216.77) |
| **Second-generation antihistamines** | 77,877,080,117<br>(857.04) | 79,719,895,342<br>(883.03) | 65,223,264,726<br>(723.75) | 62,581,875,912<br>(700.91) | 74,489,739,050<br>(839.69) | 74,742,075,676<br>(848.09) |
| **Heparinoid-containing topical preparations** | 46,029,778,556<br>(506.56) | 50,987,653,105<br>(564.77) | 52,991,914,061<br>(588.02) | 55,832,211,319<br>(625.31) | 51,956,765,121<br>(585.69) | 49,709,459,990<br>(564.05) |
| **Age 65–74 years** |  |  |  |  |  |  |
| **Prescription volume (number) of the study drugs** |  |  |  |  |  |  |
|  | **2018** | **2019** | **2020** | **2021** | **2022** | **2023** |
| **Oral single-agent acetaminophen** | 158,764,205<br>(9.02) | 171,391,962<br>(9.85) | 177,696,398<br>(10.20) | 230,764,204<br>(13.16) | 228,383,876<br>(13.54) | 263,197,595<br>(16.30) |
| **Oral single-agent NSAIDs** | 243,168,982<br>(13.81) | 233,509,002<br>(13.42) | 224,427,570<br>(12.88) | 235,428,587<br>(13.42) | 239,531,276<br>(14.20) | 242,508,351<br>(15.02) |
| **Topical anti-inflammatory patches** | 975,029,825<br>(55.39) | 953,911,163<br>(54.84) | 934,770,255<br>(53.65) | 971,050,765<br>(55.36) | 1,007,293,631<br>(59.70) | 995,849,656<br>(61.67) |
| **Second-generation antihistamines** | 333,238,654<br>(18.93) | 348,345,770<br>(20.03) | 357,139,243<br>(20.50) | 384,132,085<br>(21.90) | 484,648,899<br>(28.73) | 515,498,082<br>(31.92) |
| **Heparinoid-containing topical preparations** | 277,556,473<br>(15.77) | 305,093,212<br>(17.54) | 344,639,828<br>(19.78) | 442,066,521<br>(25.20) | 447,787,605<br>(26.54) | 481,415,766<br>(29.81) |
| **Prescription costs (Japanese Yen) of the study drugs** |  |  |  |  |  |  |
|  | **2018** | **2019** | **2020** | **2021** | **2022** | **2023** |
| **Oral single-agent acetaminophen** | 1,223,391,366<br>(69.50) | 1,334,825,072<br>(76.74) | 1,332,263,886<br>(76.46) | 1,620,705,750<br>(92.40) | 1,520,521,191<br>(90.12) | 1,715,639,272<br>(106.24) |
| **Oral single-agent NSAIDs** | 12,031,643,811<br>(683.50) | 11,772,323,880<br>(676.80) | 7,296,049,432<br>(418.71) | 4,844,290,641<br>(276.17) | 3,587,758,385<br>(212.65) | 3,068,415,321<br>(190.01) |
| **Topical anti-inflammatory patches** | 25,356,273,804<br>(1440.45) | 24,500,240,136<br>(1408.55) | 22,144,204,062<br>(1270.83) | 21,645,421,685<br>(1233.99) | 20,373,773,138<br>(1207.55) | 19,259,144,834<br>(1192.59) |
| **Second-generation antihistamines** | 15,813,921,166<br>(898.37) | 16,422,084,036<br>(944.12) | 14,403,125,993<br>(826.58) | 13,813,379,139<br>(787.49) | 14,818,851,374<br>(878.31) | 14,828,090,960<br>(918.20) |
| **Heparinoid-containing topical preparations** | 4,750,419,304<br>(269.86) | 5,117,762,373<br>(294.23) | 5,343,153,478<br>(306.64) | 5,853,281,093<br>(333.69) | 5,373,465,471<br>(318.48) | 5,206,338,956<br>(322.39) |
| **Age ≥75 years** |  |  |  |  |  |  |
| **Prescription volume (number) of the study drugs** |  |  |  |  |  |  |
|  | **2018** | **2019** | **2020** | **2021** | **2022** | **2023** |
| **Oral single-agent acetaminophen** | 451,056,598<br>(25.09) | 520,682,019<br>(28.16) | 580,351,376<br>(31.20) | 694,823,727<br>(37.21) | 726,101,725<br>(37.50) | 832,308,232<br>(41.46) |
| **Oral single-agent NSAIDs** | 491,058,149<br>(27.32) | 476,695,639<br>(25.78) | 448,551,130<br>(24.11) | 450,266,655<br>(24.11) | 474,832,121<br>(24.52) | 491,693,864<br>(24.49) |
| **Topical anti-inflammatory patches** | 2,210,694,172<br>(122.97) | 2,209,514,997<br>(119.50) | 2,112,549,019<br>(113.57) | 2,126,449,194<br>(113.88) | 2,319,574,615<br>(119.79) | 2,387,635,637<br>(118.94) |
| **Second-generation antihistamines** | 415,358,307<br>(23.10) | 442,193,464<br>(23.92) | 445,521,576<br>(23.95) | 474,878,588<br>(25.43) | 614,382,129<br>(31.73) | 679,960,291<br>(33.87) |
| **Heparinoid-containing topical preparations** | 553,227,591<br>(30.77) | 624,923,740<br>(33.80) | 691,846,826<br>(37.19) | 887,212,729<br>(47.52) | 946,255,632<br>(48.87) | 1,065,159,430<br>(53.06) |
| **Prescription costs (Japanese Yen) of the study drugs** |  |  |  |  |  |  |
|  | **2018** | **2019** | **2020** | **2021** | **2022** | **2023** |
| **Oral single-agent acetaminophen** | 3,448,249,255<br>(191.81) | 4,012,366,524<br>(217.00) | 4,280,917,420<br>(230.13) | 4,802,506,207<br>(257.20) | 4,758,812,300<br>(245.76) | 5,348,366,630<br>(266.42) |
| **Oral single-agent NSAIDs** | 26,873,322,844<br>(1494.87) | 26,632,877,419<br>(1440.39) | 16,440,846,714<br>(883.82) | 10,330,797,810<br>(553.28) | 7,644,559,657<br>(394.78) | 6,665,842,438<br>(332.05) |
| **Topical anti-inflammatory patches** | 58,148,274,994<br>(3234.59) | 57,815,783,370<br>(3126.87) | 51,300,091,532<br>(2757.77) | 48,752,835,771<br>(2611.01) | 48,010,518,016<br>(2479.37) | 47,200,583,564<br>(2351.21) |
| **Second-generation antihistamines** | 20,137,400,825<br>(1120.18) | 21,243,461,340<br>(1148.92) | 18,288,550,629<br>(983.15) | 17,328,722,039<br>(928.06) | 18,896,335,820<br>(975.85) | 19,547,915,431<br>(973.74) |
| **Heparinoid-containing topical preparations** | 9,419,041,953<br>(523.95) | 10,364,501,808<br>(560.55) | 10,563,190,156<br>(567.85) | 11,526,483,192<br>(617.31) | 11,092,523,911<br>(572.84) | 11,198,048,921<br>(557.81) |

The data in parentheses represent per capita values.
OTC: over-the-counter, NDB: National Database of Health Insurance Claims, NSAIDs: nonsteroidal anti-inflammatory drugs

**Supplementary Table 5.**
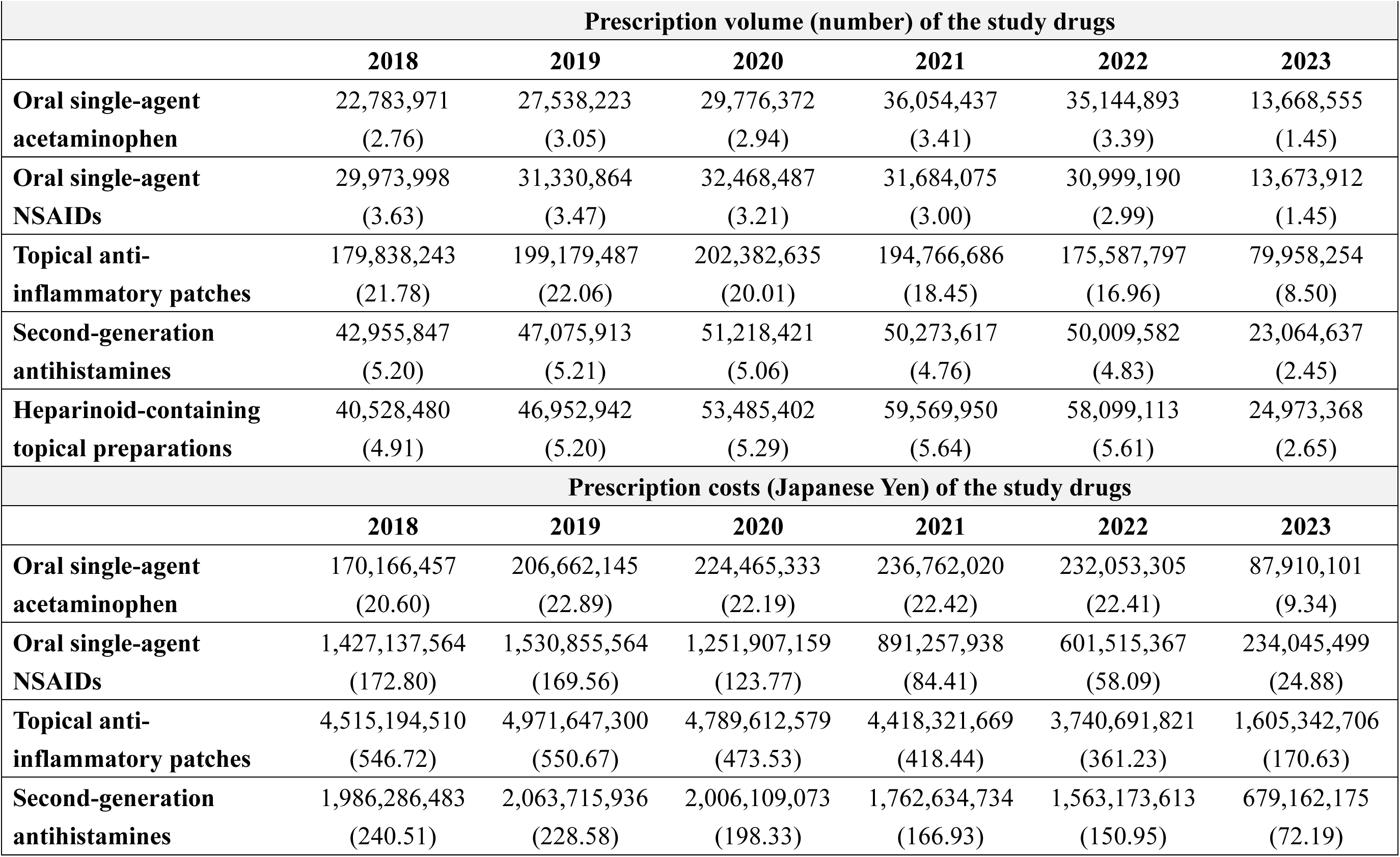
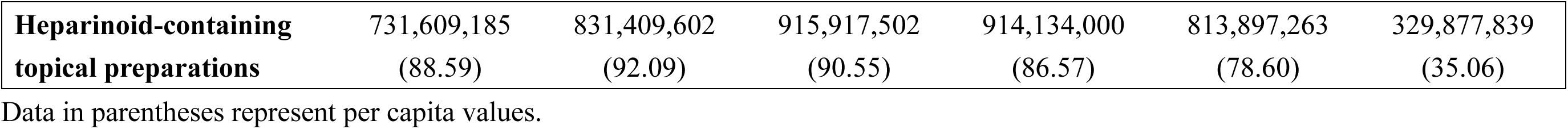
Annual prescription volume and costs of representative OTC-like drugs: Analysis of the DeSC database.

**Supplementary Table 6.**
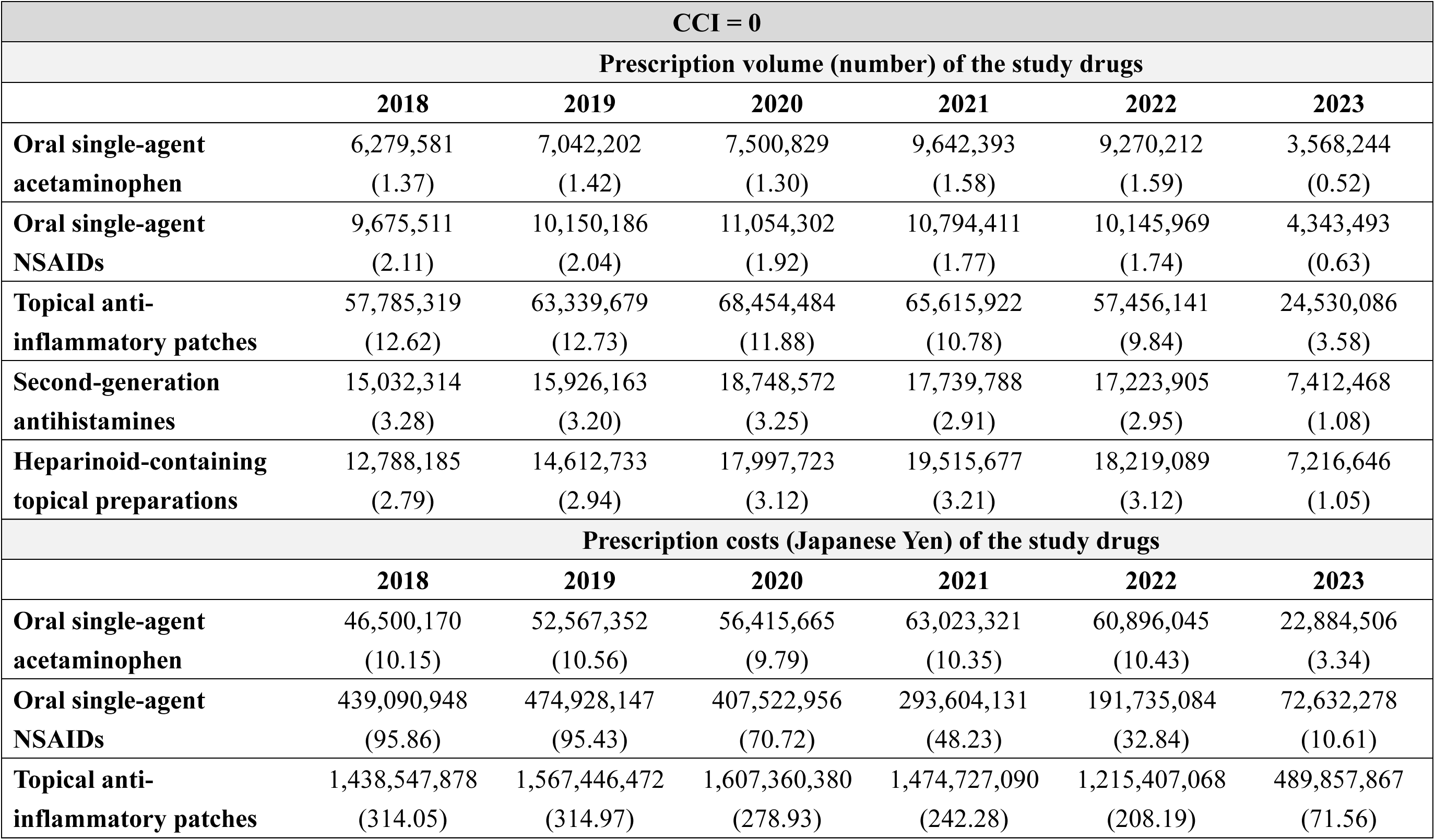
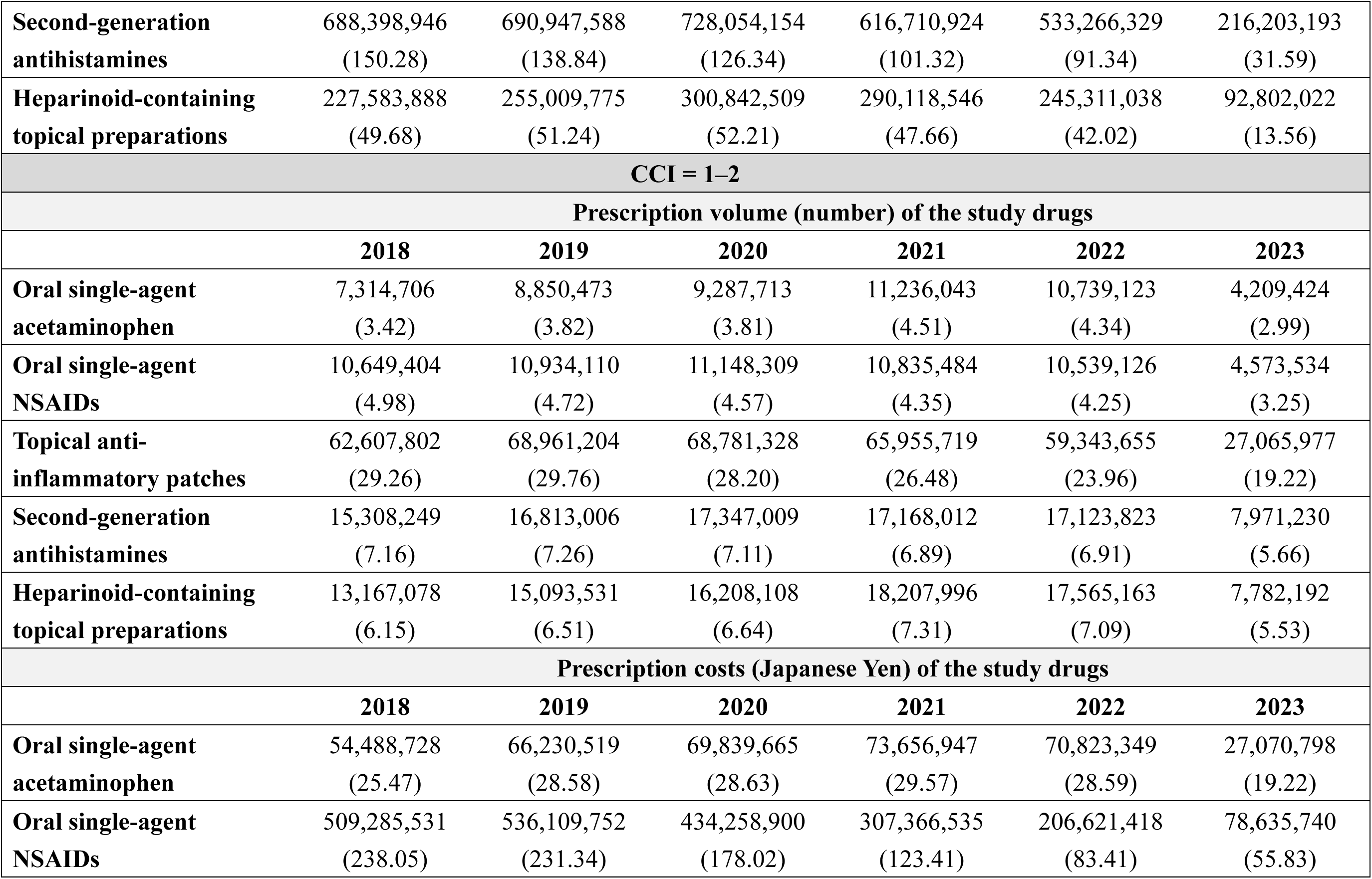
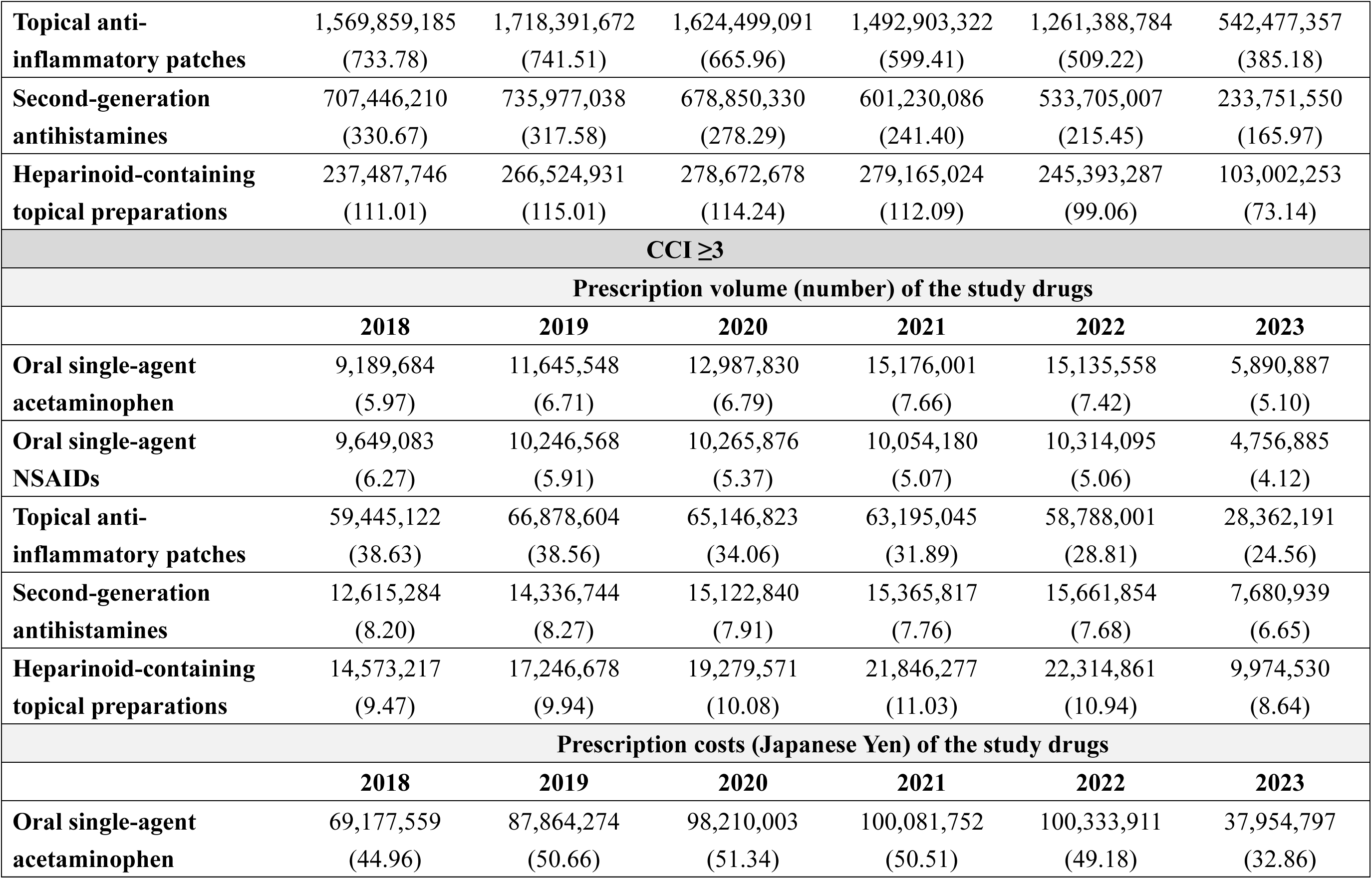
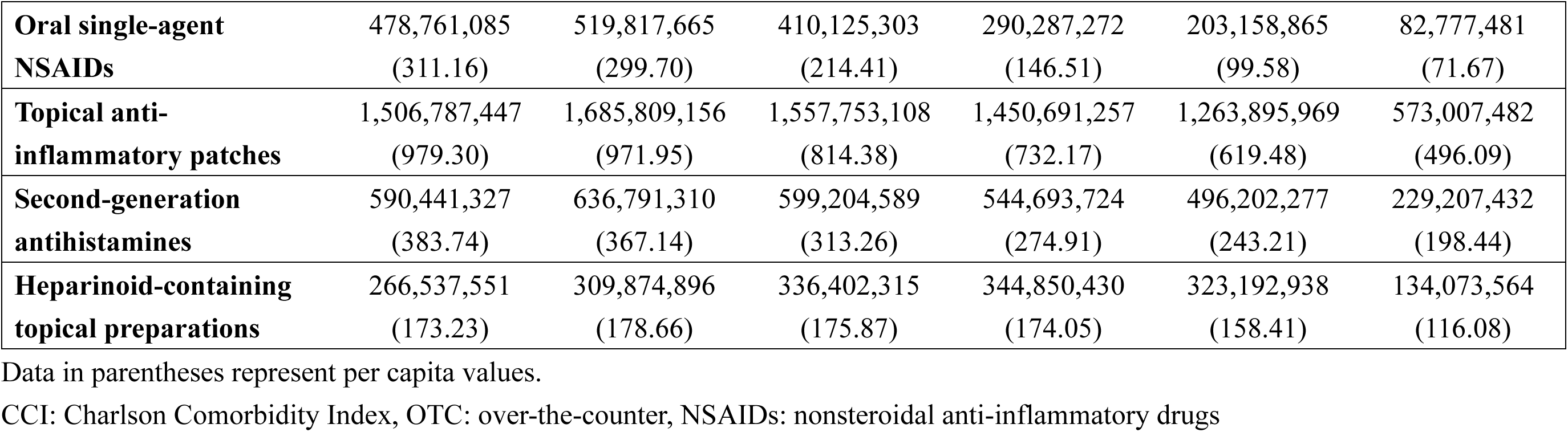
Annual prescription volume and costs for representative OTC-like drugs stratified by CCI categories: Analysis of the DeSC database.

| **CCI = 0** |  |  |  |  |  |  |
| --- | --- | --- | --- | --- | --- | --- |
| **Prescription volume (number) of the study drugs** |  |  |  |  |  |  |
|  | **2018** | **2019** | **2020** | **2021** | **2022** | **2023** |
| **Oral single-agent acetaminophen** | 6,279,581<br>(1.37) | 7,042,202<br>(1.42) | 7,500,829<br>(1.30) | 9,642,393<br>(1.58) | 9,270,212<br>(1.59) | 3,568,244<br>(0.52) |
| **Oral single-agent NSAIDs** | 9,675,511<br>(2.11) | 10,150,186<br>(2.04) | 11,054,302<br>(1.92) | 10,794,411<br>(1.77) | 10,145,969<br>(1.74) | 4,343,493<br>(0.63) |
| **Topical anti-inflammatory patches** | 57,785,319<br>(12.62) | 63,339,679<br>(12.73) | 68,454,484<br>(11.88) | 65,615,922<br>(10.78) | 57,456,141<br>(9.84) | 24,530,086<br>(3.58) |
| **Second-generation antihistamines** | 15,032,314<br>(3.28) | 15,926,163<br>(3.20) | 18,748,572<br>(3.25) | 17,739,788<br>(2.91) | 17,223,905<br>(2.95) | 7,412,468<br>(1.08) |
| **Heparinoid-containing topical preparations** | 12,788,185<br>(2.79) | 14,612,733<br>(2.94) | 17,997,723<br>(3.12) | 19,515,677<br>(3.21) | 18,219,089<br>(3.12) | 7,216,646<br>(1.05) |
| **Prescription costs (Japanese Yen) of the study drugs** |  |  |  |  |  |  |
|  | **2018** | **2019** | **2020** | **2021** | **2022** | **2023** |
| **Oral single-agent acetaminophen** | 46,500,170<br>(10.15) | 52,567,352<br>(10.56) | 56,415,665<br>(9.79) | 63,023,321<br>(10.35) | 60,896,045<br>(10.43) | 22,884,506<br>(3.34) |
| **Oral single-agent NSAIDs** | 439,090,948<br>(95.86) | 474,928,147<br>(95.43) | 407,522,956<br>(70.72) | 293,604,131<br>(48.23) | 191,735,084<br>(32.84) | 72,632,278<br>(10.61) |
| **Topical anti-inflammatory patches** | 1,438,547,878<br>(314.05) | 1,567,446,472<br>(314.97) | 1,607,360,380<br>(278.93) | 1,474,727,090<br>(242.28) | 1,215,407,068<br>(208.19) | 489,857,867<br>(71.56) |
| **Second-generation antihistamines** | 688,398,946<br>(150.28) | 690,947,588<br>(138.84) | 728,054,154<br>(126.34) | 616,710,924<br>(101.32) | 533,266,329<br>(91.34) | 216,203,193<br>(31.59) |
| **Heparinoid-containing topical preparations** | 227,583,888<br>(49.68) | 255,009,775<br>(51.24) | 300,842,509<br>(52.21) | 290,118,546<br>(47.66) | 245,311,038<br>(42.02) | 92,802,022<br>(13.56) |
| **CCI = 1–2** |  |  |  |  |  |  |
| **Prescription volume (number) of the study drugs** |  |  |  |  |  |  |
|  | **2018** | **2019** | **2020** | **2021** | **2022** | **2023** |
| **Oral single-agent acetaminophen** | 7,314,706<br>(3.42) | 8,850,473<br>(3.82) | 9,287,713<br>(3.81) | 11,236,043<br>(4.51) | 10,739,123<br>(4.34) | 4,209,424<br>(2.99) |
| **Oral single-agent NSAIDs** | 10,649,404<br>(4.98) | 10,934,110<br>(4.72) | 11,148,309<br>(4.57) | 10,835,484<br>(4.35) | 10,539,126<br>(4.25) | 4,573,534<br>(3.25) |
| **Topical anti-inflammatory patches** | 62,607,802<br>(29.26) | 68,961,204<br>(29.76) | 68,781,328<br>(28.20) | 65,955,719<br>(26.48) | 59,343,655<br>(23.96) | 27,065,977<br>(19.22) |
| **Second-generation antihistamines** | 15,308,249<br>(7.16) | 16,813,006<br>(7.26) | 17,347,009<br>(7.11) | 17,168,012<br>(6.89) | 17,123,823<br>(6.91) | 7,971,230<br>(5.66) |
| **Heparinoid-containing topical preparations** | 13,167,078<br>(6.15) | 15,093,531<br>(6.51) | 16,208,108<br>(6.64) | 18,207,996<br>(7.31) | 17,565,163<br>(7.09) | 7,782,192<br>(5.53) |
| **Prescription costs (Japanese Yen) of the study drugs** |  |  |  |  |  |  |
|  | **2018** | **2019** | **2020** | **2021** | **2022** | **2023** |
| **Oral single-agent acetaminophen** | 54,488,728<br>(25.47) | 66,230,519<br>(28.58) | 69,839,665<br>(28.63) | 73,656,947<br>(29.57) | 70,823,349<br>(28.59) | 27,070,798<br>(19.22) |
| **Oral single-agent NSAIDs** | 509,285,531<br>(238.05) | 536,109,752<br>(231.34) | 434,258,900<br>(178.02) | 307,366,535<br>(123.41) | 206,621,418<br>(83.41) | 78,635,740<br>(55.83) |
| **Topical anti-inflammatory patches** | 1,569,859,185<br>(733.78) | 1,718,391,672<br>(741.51) | 1,624,499,091<br>(665.96) | 1,492,903,322<br>(599.41) | 1,261,388,784<br>(509.22) | 542,477,357<br>(385.18) |
| **Second-generation antihistamines** | 707,446,210<br>(330.67) | 735,977,038<br>(317.58) | 678,850,330<br>(278.29) | 601,230,086<br>(241.40) | 533,705,007<br>(215.45) | 233,751,550<br>(165.97) |
| **Heparinoid-containing topical preparations** | 237,487,746<br>(111.01) | 266,524,931<br>(115.01) | 278,672,678<br>(114.24) | 279,165,024<br>(112.09) | 245,393,287<br>(99.06) | 103,002,253<br>(73.14) |
| **CCI ≥3** |  |  |  |  |  |  |
| **Prescription volume (number) of the study drugs** |  |  |  |  |  |  |
|  | **2018** | **2019** | **2020** | **2021** | **2022** | **2023** |
| **Oral single-agent acetaminophen** | 9,189,684<br>(5.97) | 11,645,548<br>(6.71) | 12,987,830<br>(6.79) | 15,176,001<br>(7.66) | 15,135,558<br>(7.42) | 5,890,887<br>(5.10) |
| **Oral single-agent NSAIDs** | 9,649,083<br>(6.27) | 10,246,568<br>(5.91) | 10,265,876<br>(5.37) | 10,054,180<br>(5.07) | 10,314,095<br>(5.06) | 4,756,885<br>(4.12) |
| **Topical anti-inflammatory patches** | 59,445,122<br>(38.63) | 66,878,604<br>(38.56) | 65,146,823<br>(34.06) | 63,195,045<br>(31.89) | 58,788,001<br>(28.81) | 28,362,191<br>(24.56) |
| **Second-generation antihistamines** | 12,615,284<br>(8.20) | 14,336,744<br>(8.27) | 15,122,840<br>(7.91) | 15,365,817<br>(7.76) | 15,661,854<br>(7.68) | 7,680,939<br>(6.65) |
| **Heparinoid-containing topical preparations** | 14,573,217<br>(9.47) | 17,246,678<br>(9.94) | 19,279,571<br>(10.08) | 21,846,277<br>(11.03) | 22,314,861<br>(10.94) | 9,974,530<br>(8.64) |
| **Prescription costs (Japanese Yen) of the study drugs** |  |  |  |  |  |  |
|  | **2018** | **2019** | **2020** | **2021** | **2022** | **2023** |
| **Oral single-agent acetaminophen** | 69,177,559<br>(44.96) | 87,864,274<br>(50.66) | 98,210,003<br>(51.34) | 100,081,752<br>(50.51) | 100,333,911<br>(49.18) | 37,954,797<br>(32.86) |
| **Oral single-agent NSAIDs** | 478,761,085<br>(311.16) | 519,817,665<br>(299.70) | 410,125,303<br>(214.41) | 290,287,272<br>(146.51) | 203,158,865<br>(99.58) | 82,777,481<br>(71.67) |
| **Topical anti-inflammatory patches** | 1,506,787,447<br>(979.30) | 1,685,809,156<br>(971.95) | 1,557,753,108<br>(814.38) | 1,450,691,257<br>(732.17) | 1,263,895,969<br>(619.48) | 573,007,482<br>(496.09) |
| **Second-generation antihistamines** | 590,441,327<br>(383.74) | 636,791,310<br>(367.14) | 599,204,589<br>(313.26) | 544,693,724<br>(274.91) | 496,202,277<br>(243.21) | 229,207,432<br>(198.44) |
| **Heparinoid-containing topical preparations** | 266,537,551<br>(173.23) | 309,874,896<br>(178.66) | 336,402,315<br>(175.87) | 344,850,430<br>(174.05) | 323,192,938<br>(158.41) | 134,073,564<br>(116.08) |

Data in parentheses represent per capita values.
CCI: Charlson Comorbidity Index, OTC: over-the-counter, NSAIDs: nonsteroidal anti-inflammatory drugs

**Supplementary Table 7.**
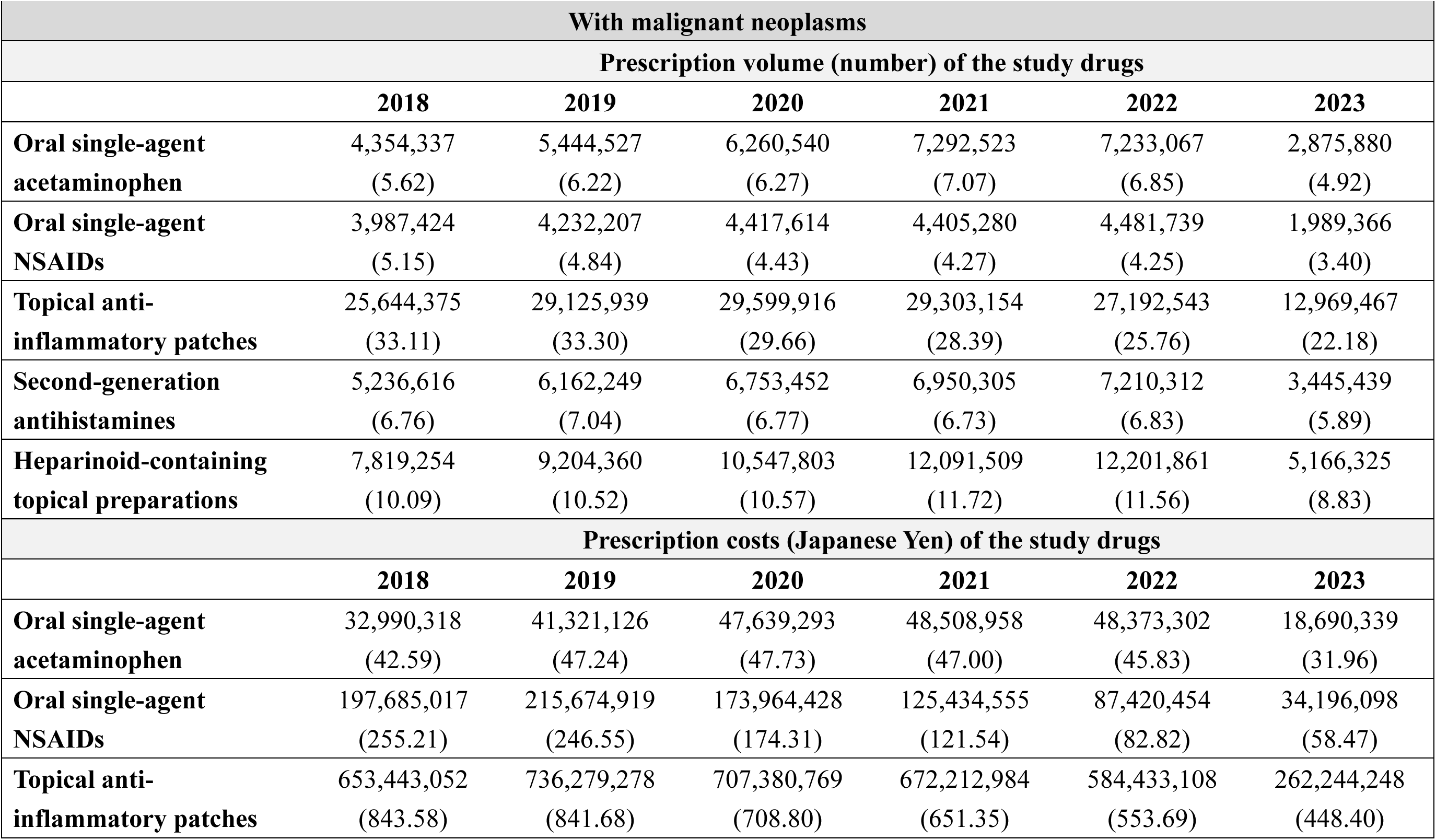
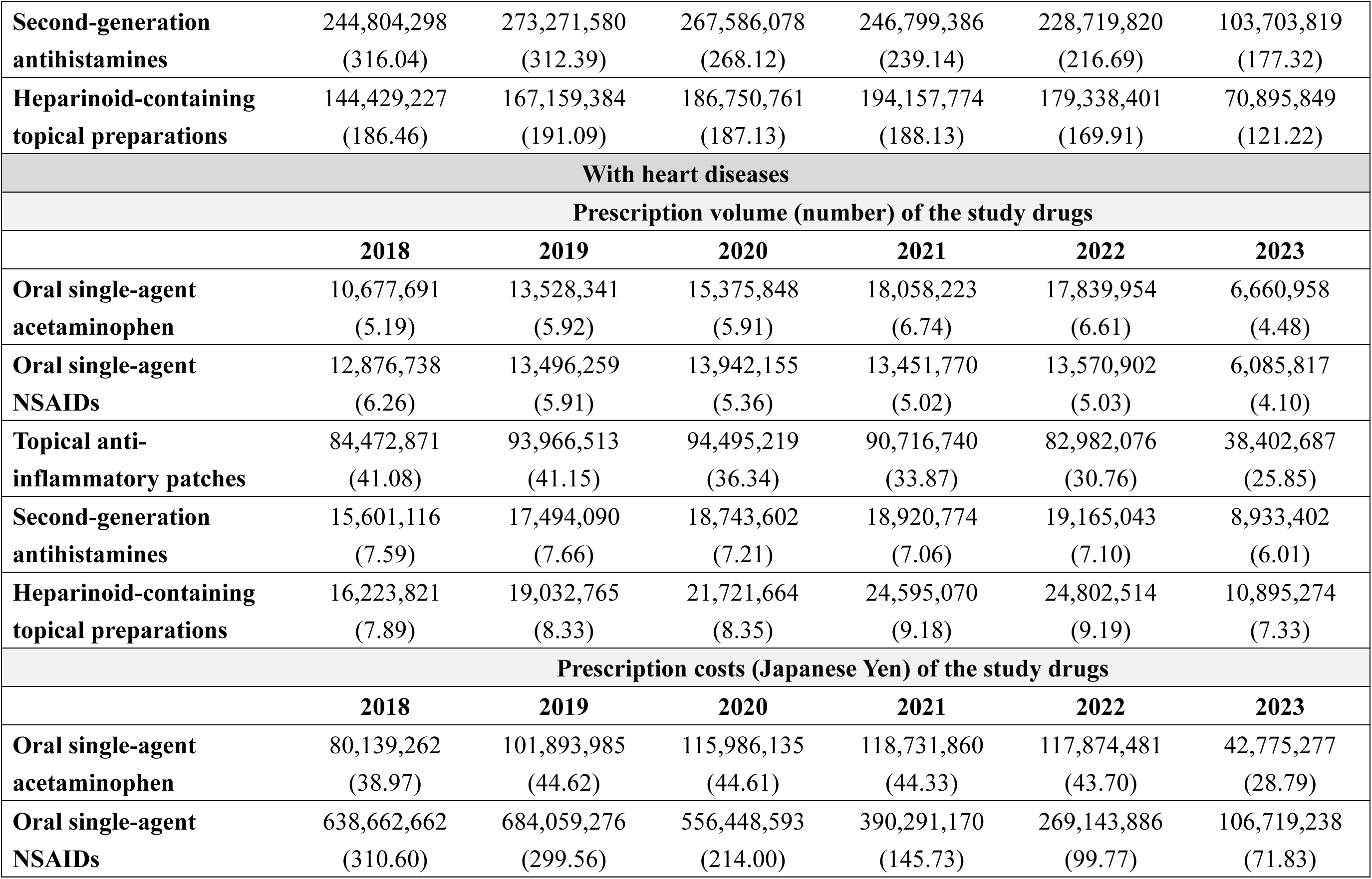
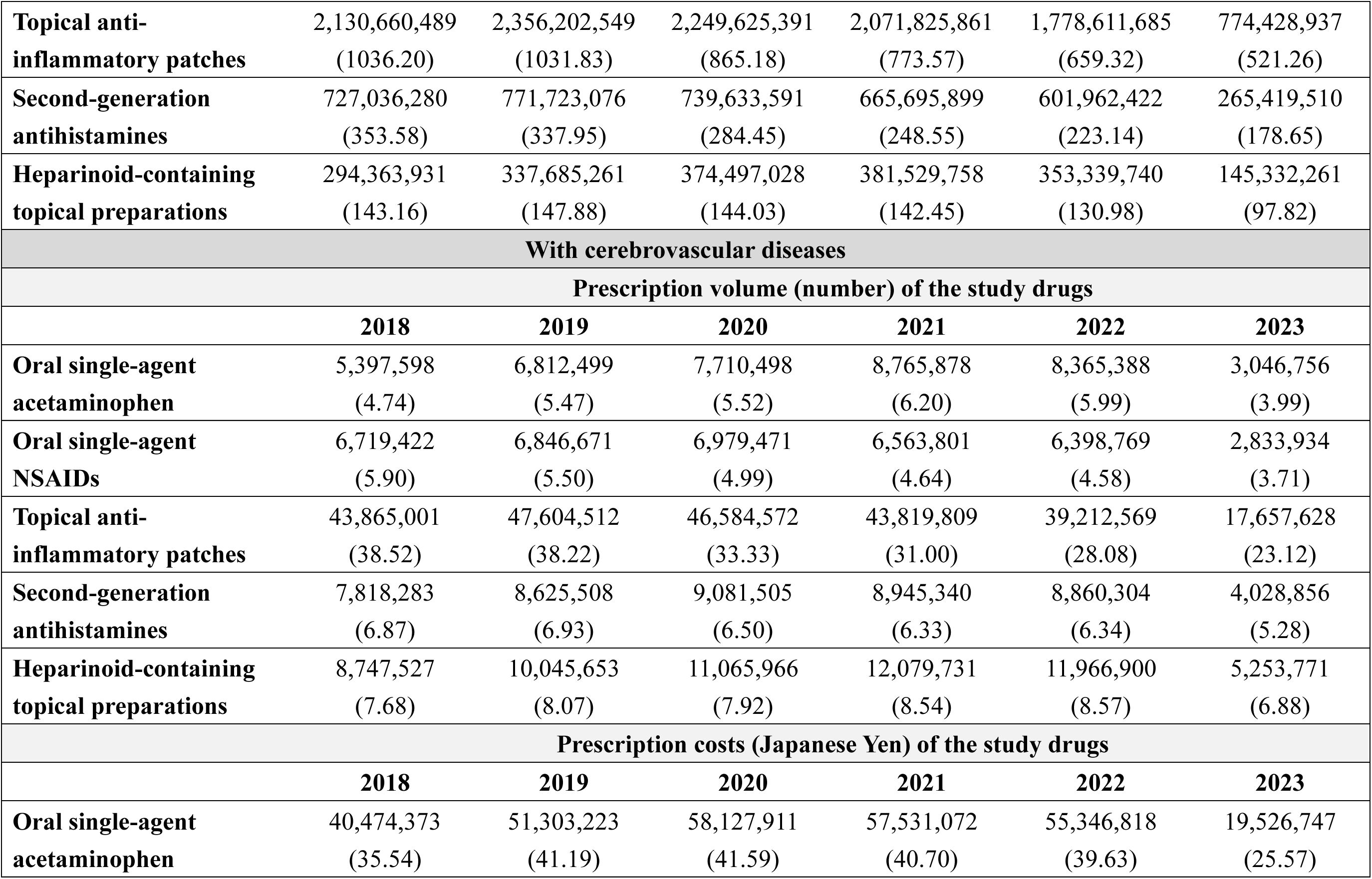
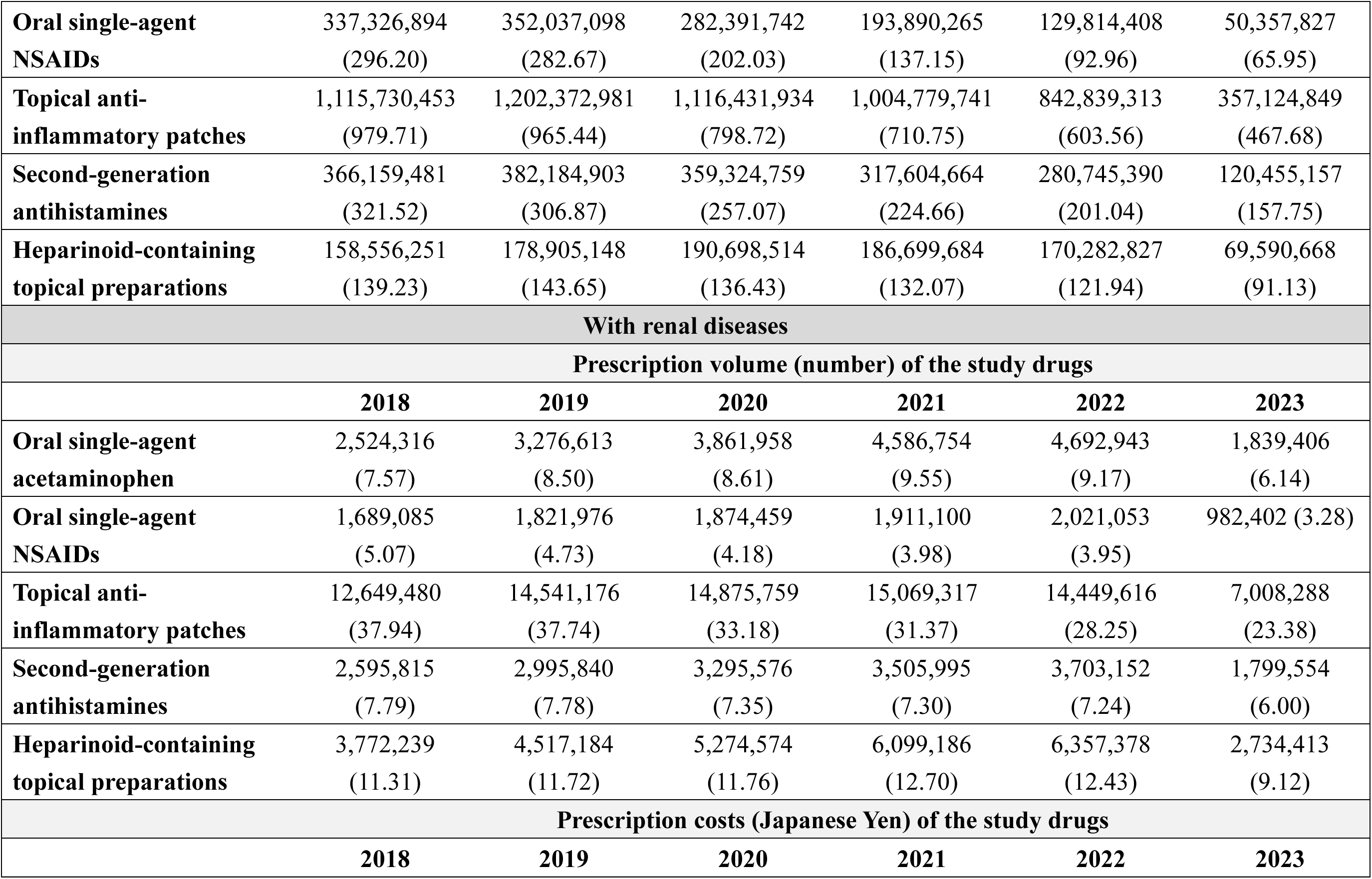
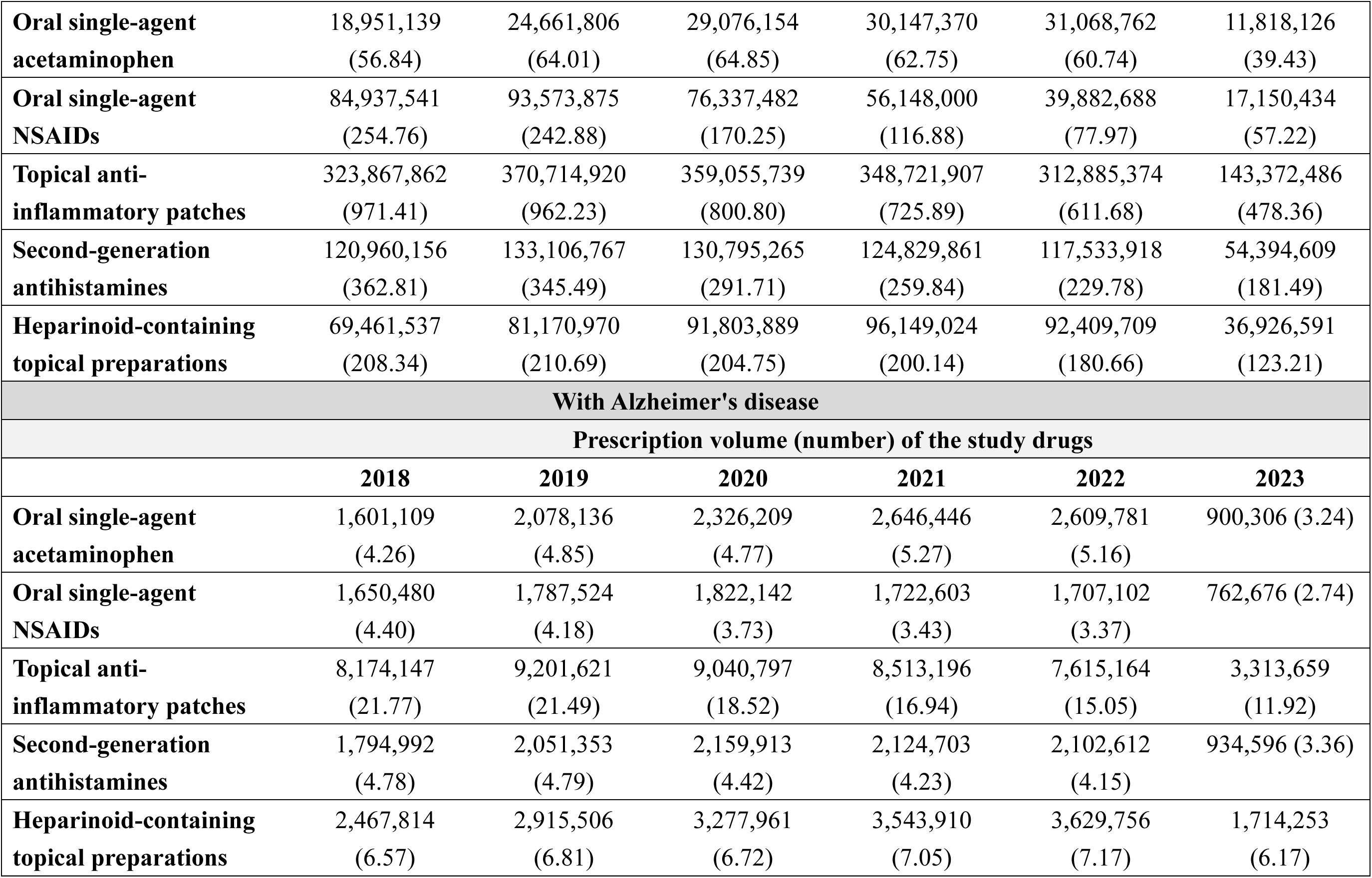
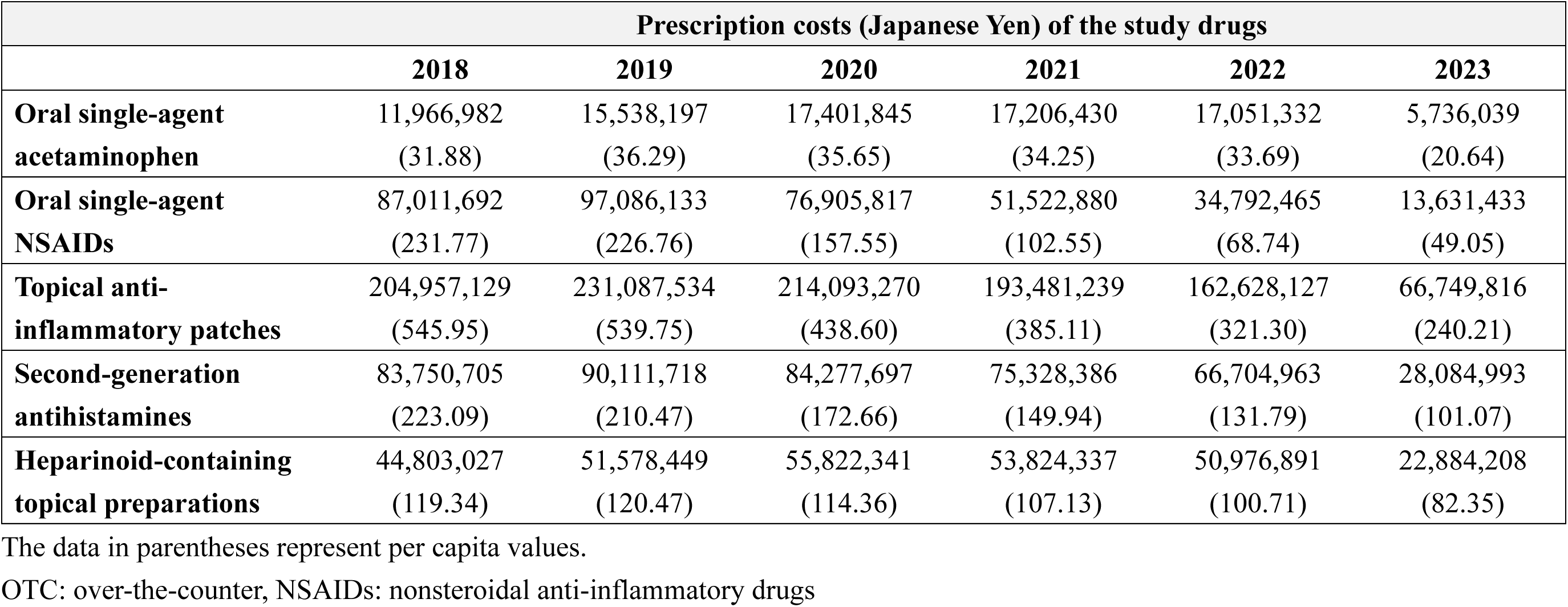
Annual prescription volume and costs of representative OTC-like drugs by the presence of Japan’s leading causes of death: Analysis of the DeSC database.

| **With malignant neoplasms** |  |  |  |  |  |  |
| --- | --- | --- | --- | --- | --- | --- |
| **Prescription volume (number) of the study drugs** |  |  |  |  |  |  |
|  | **2018** | **2019** | **2020** | **2021** | **2022** | **2023** |
| **Oral single-agent acetaminophen** | 4,354,337<br>(5.62) | 5,444,527<br>(6.22) | 6,260,540<br>(6.27) | 7,292,523<br>(7.07) | 7,233,067<br>(6.85) | 2,875,880<br>(4.92) |
| **Oral single-agent NSAIDs** | 3,987,424<br>(5.15) | 4,232,207<br>(4.84) | 4,417,614<br>(4.43) | 4,405,280<br>(4.27) | 4,481,739<br>(4.25) | 1,989,366<br>(3.40) |
| **Topical anti-inflammatory patches** | 25,644,375<br>(33.11) | 29,125,939<br>(33.30) | 29,599,916<br>(29.66) | 29,303,154<br>(28.39) | 27,192,543<br>(25.76) | 12,969,467<br>(22.18) |
| **Second-generation antihistamines** | 5,236,616<br>(6.76) | 6,162,249<br>(7.04) | 6,753,452<br>(6.77) | 6,950,305<br>(6.73) | 7,210,312<br>(6.83) | 3,445,439<br>(5.89) |
| **Heparinoid-containing topical preparations** | 7,819,254<br>(10.09) | 9,204,360<br>(10.52) | 10,547,803<br>(10.57) | 12,091,509<br>(11.72) | 12,201,861<br>(11.56) | 5,166,325<br>(8.83) |
| **Prescription costs (Japanese Yen) of the study drugs** |  |  |  |  |  |  |
|  | **2018** | **2019** | **2020** | **2021** | **2022** | **2023** |
| **Oral single-agent acetaminophen** | 32,990,318<br>(42.59) | 41,321,126<br>(47.24) | 47,639,293<br>(47.73) | 48,508,958<br>(47.00) | 48,373,302<br>(45.83) | 18,690,339<br>(31.96) |
| **Oral single-agent NSAIDs** | 197,685,017<br>(255.21) | 215,674,919<br>(246.55) | 173,964,428<br>(174.31) | 125,434,555<br>(121.54) | 87,420,454<br>(82.82) | 34,196,098<br>(58.47) |
| **Topical anti-inflammatory patches** | 653,443,052<br>(843.58) | 736,279,278<br>(841.68) | 707,380,769<br>(708.80) | 672,212,984<br>(651.35) | 584,433,108<br>(553.69) | 262,244,248<br>(448.40) |
| **Second-generation antihistamines** | 244,804,298<br>(316.04) | 273,271,580<br>(312.39) | 267,586,078<br>(268.12) | 246,799,386<br>(239.14) | 228,719,820<br>(216.69) | 103,703,819<br>(177.32) |
| **Heparinoid-containing topical preparations** | 144,429,227<br>(186.46) | 167,159,384<br>(191.09) | 186,750,761<br>(187.13) | 194,157,774<br>(188.13) | 179,338,401<br>(169.91) | 70,895,849<br>(121.22) |
| **With heart diseases** |  |  |  |  |  |  |
| **Prescription volume (number) of the study drugs** |  |  |  |  |  |  |
|  | **2018** | **2019** | **2020** | **2021** | **2022** | **2023** |
| **Oral single-agent acetaminophen** | 10,677,691<br>(5.19) | 13,528,341<br>(5.92) | 15,375,848<br>(5.91) | 18,058,223<br>(6.74) | 17,839,954<br>(6.61) | 6,660,958<br>(4.48) |
| **Oral single-agent NSAIDs** | 12,876,738<br>(6.26) | 13,496,259<br>(5.91) | 13,942,155<br>(5.36) | 13,451,770<br>(5.02) | 13,570,902<br>(5.03) | 6,085,817<br>(4.10) |
| **Topical anti-inflammatory patches** | 84,472,871<br>(41.08) | 93,966,513<br>(41.15) | 94,495,219<br>(36.34) | 90,716,740<br>(33.87) | 82,982,076<br>(30.76) | 38,402,687<br>(25.85) |
| **Second-generation antihistamines** | 15,601,116<br>(7.59) | 17,494,090<br>(7.66) | 18,743,602<br>(7.21) | 18,920,774<br>(7.06) | 19,165,043<br>(7.10) | 8,933,402<br>(6.01) |
| **Heparinoid-containing topical preparations** | 16,223,821<br>(7.89) | 19,032,765<br>(8.33) | 21,721,664<br>(8.35) | 24,595,070<br>(9.18) | 24,802,514<br>(9.19) | 10,895,274<br>(7.33) |
| **Prescription costs (Japanese Yen) of the study drugs** |  |  |  |  |  |  |
|  | **2018** | **2019** | **2020** | **2021** | **2022** | **2023** |
| **Oral single-agent acetaminophen** | 80,139,262<br>(38.97) | 101,893,985<br>(44.62) | 115,986,135<br>(44.61) | 118,731,860<br>(44.33) | 117,874,481<br>(43.70) | 42,775,277<br>(28.79) |
| **Oral single-agent NSAIDs** | 638,662,662<br>(310.60) | 684,059,276<br>(299.56) | 556,448,593<br>(214.00) | 390,291,170<br>(145.73) | 269,143,886<br>(99.77) | 106,719,238<br>(71.83) |
| **Topical anti-inflammatory patches** | 2,130,660,489<br>(1036.20) | 2,356,202,549<br>(1031.83) | 2,249,625,391<br>(865.18) | 2,071,825,861<br>(773.57) | 1,778,611,685<br>(659.32) | 774,428,937<br>(521.26) |
| **Second-generation antihistamines** | 727,036,280<br>(353.58) | 771,723,076<br>(337.95) | 739,633,591<br>(284.45) | 665,695,899<br>(248.55) | 601,962,422<br>(223.14) | 265,419,510<br>(178.65) |
| **Heparinoid-containing topical preparations** | 294,363,931<br>(143.16) | 337,685,261<br>(147.88) | 374,497,028<br>(144.03) | 381,529,758<br>(142.45) | 353,339,740<br>(130.98) | 145,332,261<br>(97.82) |
| **With cerebrovascular diseases** |  |  |  |  |  |  |
| **Prescription volume (number) of the study drugs** |  |  |  |  |  |  |
|  | **2018** | **2019** | **2020** | **2021** | **2022** | **2023** |
| **Oral single-agent acetaminophen** | 5,397,598<br>(4.74) | 6,812,499<br>(5.47) | 7,710,498<br>(5.52) | 8,765,878<br>(6.20) | 8,365,388<br>(5.99) | 3,046,756<br>(3.99) |
| **Oral single-agent NSAIDs** | 6,719,422<br>(5.90) | 6,846,671<br>(5.50) | 6,979,471<br>(4.99) | 6,563,801<br>(4.64) | 6,398,769<br>(4.58) | 2,833,934<br>(3.71) |
| **Topical anti-inflammatory patches** | 43,865,001<br>(38.52) | 47,604,512<br>(38.22) | 46,584,572<br>(33.33) | 43,819,809<br>(31.00) | 39,212,569<br>(28.08) | 17,657,628<br>(23.12) |
| **Second-generation antihistamines** | 7,818,283<br>(6.87) | 8,625,508<br>(6.93) | 9,081,505<br>(6.50) | 8,945,340<br>(6.33) | 8,860,304<br>(6.34) | 4,028,856<br>(5.28) |
| **Heparinoid-containing topical preparations** | 8,747,527<br>(7.68) | 10,045,653<br>(8.07) | 11,065,966<br>(7.92) | 12,079,731<br>(8.54) | 11,966,900<br>(8.57) | 5,253,771<br>(6.88) |
| **Prescription costs (Japanese Yen) of the study drugs** |  |  |  |  |  |  |
|  | **2018** | **2019** | **2020** | **2021** | **2022** | **2023** |
| **Oral single-agent acetaminophen** | 40,474,373<br>(35.54) | 51,303,223<br>(41.19) | 58,127,911<br>(41.59) | 57,531,072<br>(40.70) | 55,346,818<br>(39.63) | 19,526,747<br>(25.57) |
| **Oral single-agent NSAIDs** | 337,326,894<br>(296.20) | 352,037,098<br>(282.67) | 282,391,742<br>(202.03) | 193,890,265<br>(137.15) | 129,814,408<br>(92.96) | 50,357,827<br>(65.95) |
| **Topical anti-inflammatory patches** | 1,115,730,453<br>(979.71) | 1,202,372,981<br>(965.44) | 1,116,431,934<br>(798.72) | 1,004,779,741<br>(710.75) | 842,839,313<br>(603.56) | 357,124,849<br>(467.68) |
| **Second-generation antihistamines** | 366,159,481<br>(321.52) | 382,184,903<br>(306.87) | 359,324,759<br>(257.07) | 317,604,664<br>(224.66) | 280,745,390<br>(201.04) | 120,455,157<br>(157.75) |
| **Heparinoid-containing topical preparations** | 158,556,251<br>(139.23) | 178,905,148<br>(143.65) | 190,698,514<br>(136.43) | 186,699,684<br>(132.07) | 170,282,827<br>(121.94) | 69,590,668<br>(91.13) |
| **With renal diseases** |  |  |  |  |  |  |
| **Prescription volume (number) of the study drugs** |  |  |  |  |  |  |
|  | **2018** | **2019** | **2020** | **2021** | **2022** | **2023** |
| **Oral single-agent acetaminophen** | 2,524,316<br>(7.57) | 3,276,613<br>(8.50) | 3,861,958<br>(8.61) | 4,586,754<br>(9.55) | 4,692,943<br>(9.17) | 1,839,406<br>(6.14) |
| **Oral single-agent NSAIDs** | 1,689,085<br>(5.07) | 1,821,976<br>(4.73) | 1,874,459<br>(4.18) | 1,911,100<br>(3.98) | 2,021,053<br>(3.95) | 982,402 (3.28) |
| **Topical anti-inflammatory patches** | 12,649,480<br>(37.94) | 14,541,176<br>(37.74) | 14,875,759<br>(33.18) | 15,069,317<br>(31.37) | 14,449,616<br>(28.25) | 7,008,288<br>(23.38) |
| **Second-generation antihistamines** | 2,595,815<br>(7.79) | 2,995,840<br>(7.78) | 3,295,576<br>(7.35) | 3,505,995<br>(7.30) | 3,703,152<br>(7.24) | 1,799,554<br>(6.00) |
| **Heparinoid-containing topical preparations** | 3,772,239<br>(11.31) | 4,517,184<br>(11.72) | 5,274,574<br>(11.76) | 6,099,186<br>(12.70) | 6,357,378<br>(12.43) | 2,734,413<br>(9.12) |
| **Prescription costs (Japanese Yen) of the study drugs** |  |  |  |  |  |  |
|  | **2018** | **2019** | **2020** | **2021** | **2022** | **2023** |
| **Oral single-agent acetaminophen** | 18,951,139<br>(56.84) | 24,661,806<br>(64.01) | 29,076,154<br>(64.85) | 30,147,370<br>(62.75) | 31,068,762<br>(60.74) | 11,818,126<br>(39.43) |
| **Oral single-agent NSAIDs** | 84,937,541<br>(254.76) | 93,573,875<br>(242.88) | 76,337,482<br>(170.25) | 56,148,000<br>(116.88) | 39,882,688<br>(77.97) | 17,150,434<br>(57.22) |
| **Topical anti-inflammatory patches** | 323,867,862<br>(971.41) | 370,714,920<br>(962.23) | 359,055,739<br>(800.80) | 348,721,907<br>(725.89) | 312,885,374<br>(611.68) | 143,372,486<br>(478.36) |
| **Second-generation antihistamines** | 120,960,156<br>(362.81) | 133,106,767<br>(345.49) | 130,795,265<br>(291.71) | 124,829,861<br>(259.84) | 117,533,918<br>(229.78) | 54,394,609<br>(181.49) |
| **Heparinoid-containing topical preparations** | 69,461,537<br>(208.34) | 81,170,970<br>(210.69) | 91,803,889<br>(204.75) | 96,149,024<br>(200.14) | 92,409,709<br>(180.66) | 36,926,591<br>(123.21) |
| **With Alzheimer's disease** |  |  |  |  |  |  |
| **Prescription volume (number) of the study drugs** |  |  |  |  |  |  |
|  | **2018** | **2019** | **2020** | **2021** | **2022** | **2023** |
| **Oral single-agent acetaminophen** | 1,601,109<br>(4.26) | 2,078,136<br>(4.85) | 2,326,209<br>(4.77) | 2,646,446<br>(5.27) | 2,609,781<br>(5.16) | 900,306 (3.24) |
| **Oral single-agent NSAIDs** | 1,650,480<br>(4.40) | 1,787,524<br>(4.18) | 1,822,142<br>(3.73) | 1,722,603<br>(3.43) | 1,707,102<br>(3.37) | 762,676 (2.74) |
| **Topical anti-inflammatory patches** | 8,174,147<br>(21.77) | 9,201,621<br>(21.49) | 9,040,797<br>(18.52) | 8,513,196<br>(16.94) | 7,615,164<br>(15.05) | 3,313,659<br>(11.92) |
| **Second-generation antihistamines** | 1,794,992<br>(4.78) | 2,051,353<br>(4.79) | 2,159,913<br>(4.42) | 2,124,703<br>(4.23) | 2,102,612<br>(4.15) | 934,596 (3.36) |
| **Heparinoid-containing topical preparations** | 2,467,814<br>(6.57) | 2,915,506<br>(6.81) | 3,277,961<br>(6.72) | 3,543,910<br>(7.05) | 3,629,756<br>(7.17) | 1,714,253<br>(6.17) |

| Prescription costs (Japanese Yen) of the study drugs |  |  |  |  |  |  |
| --- | --- | --- | --- | --- | --- | --- |
|  | 2018 | 2019 | 2020 | 2021 | 2022 | 2023 |
| **Oral single-agent acetaminophen** | 11,966,982<br>(31.88) | 15,538,197<br>(36.29) | 17,401,845<br>(35.65) | 17,206,430<br>(34.25) | 17,051,332<br>(33.69) | 5,736,039<br>(20.64) |
| **Oral single-agent NSAIDs** | 87,011,692<br>(231.77) | 97,086,133<br>(226.76) | 76,905,817<br>(157.55) | 51,522,880<br>(102.55) | 34,792,465<br>(68.74) | 13,631,433<br>(49.05) |
| **Topical anti-inflammatory patches** | 204,957,129<br>(545.95) | 231,087,534<br>(539.75) | 214,093,270<br>(438.60) | 193,481,239<br>(385.11) | 162,628,127<br>(321.30) | 66,749,816<br>(240.21) |
| **Second-generation antihistamines** | 83,750,705<br>(223.09) | 90,111,718<br>(210.47) | 84,277,697<br>(172.66) | 75,328,386<br>(149.94) | 66,704,963<br>(131.79) | 28,084,993<br>(101.07) |
| **Heparinoid-containing topical preparations** | 44,803,027<br>(119.34) | 51,578,449<br>(120.47) | 55,822,341<br>(114.36) | 53,824,337<br>(107.13) | 50,976,891<br>(100.71) | 22,884,208<br>(82.35) |

The data in parentheses represent per capita values.
OTC: over-the-counter, NSAIDs: nonsteroidal anti-inflammatory drugs

**Supplementary Table 8.** Number of individuals categorized by health insurance type in the DeSC database.

|  | **2018** | **2019** | **2020** | **2021** | **2022** | **2023** |
| --- | --- | --- | --- | --- | --- | --- |
| **Total** | 8,258,708<br>(100.0%) | 9,028,436<br>(100.0%) | 10,114,795<br>(100.0%) | 10,558,971<br>(100.0%) | 10,355,350<br>(100.0%) | 9,408,449<br>(100.0%) |
| **National Health Insurance** | 4,323,938<br>(52.4%) | 4,688,678<br>(51.9%) | 5,213,161<br>(51.5%) | 5,521,430<br>(52.3%) | 5,165,407<br>(49.9%) | 4,301,933<br>(45.7%) |
| **Employees' health insurance** | 854,938<br>(10.4%) | 845,090<br>(9.4%) | 840,520<br>(8.3%) | 838,140<br>(7.9%) | 829,945<br>(8.0%) | 662,029<br>(7.0%) |
| **Medical Care System for the Advanced Elderly** | 3,079,832<br>(37.3%) | 3,494,668<br>(38.7%) | 4,061,114<br>(40.2%) | 4,199,401<br>(39.8%) | 4,359,998<br>(42.1%) | 4,444,487<br>(47.2%) |

**Supplementary Table 9.**
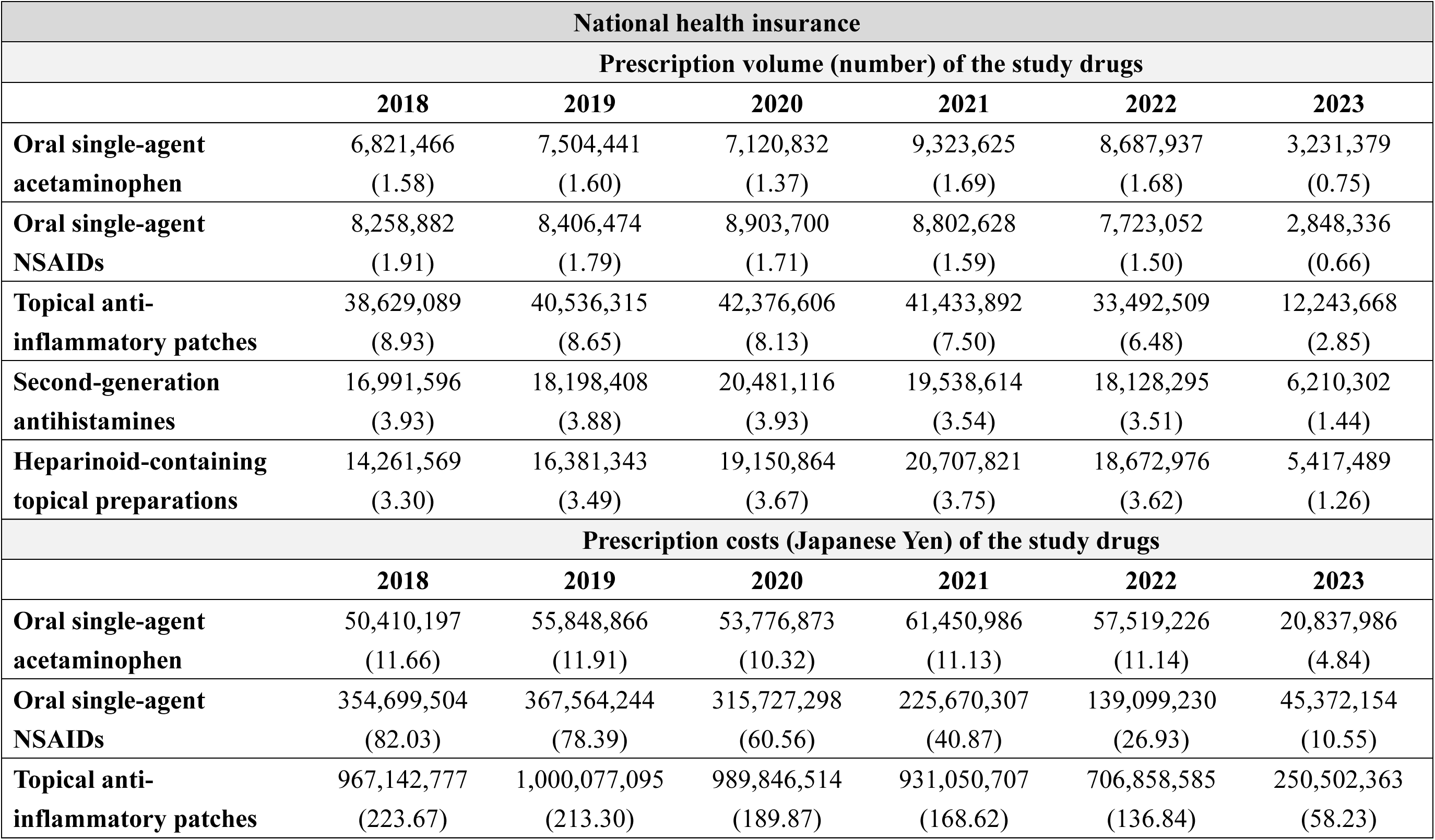
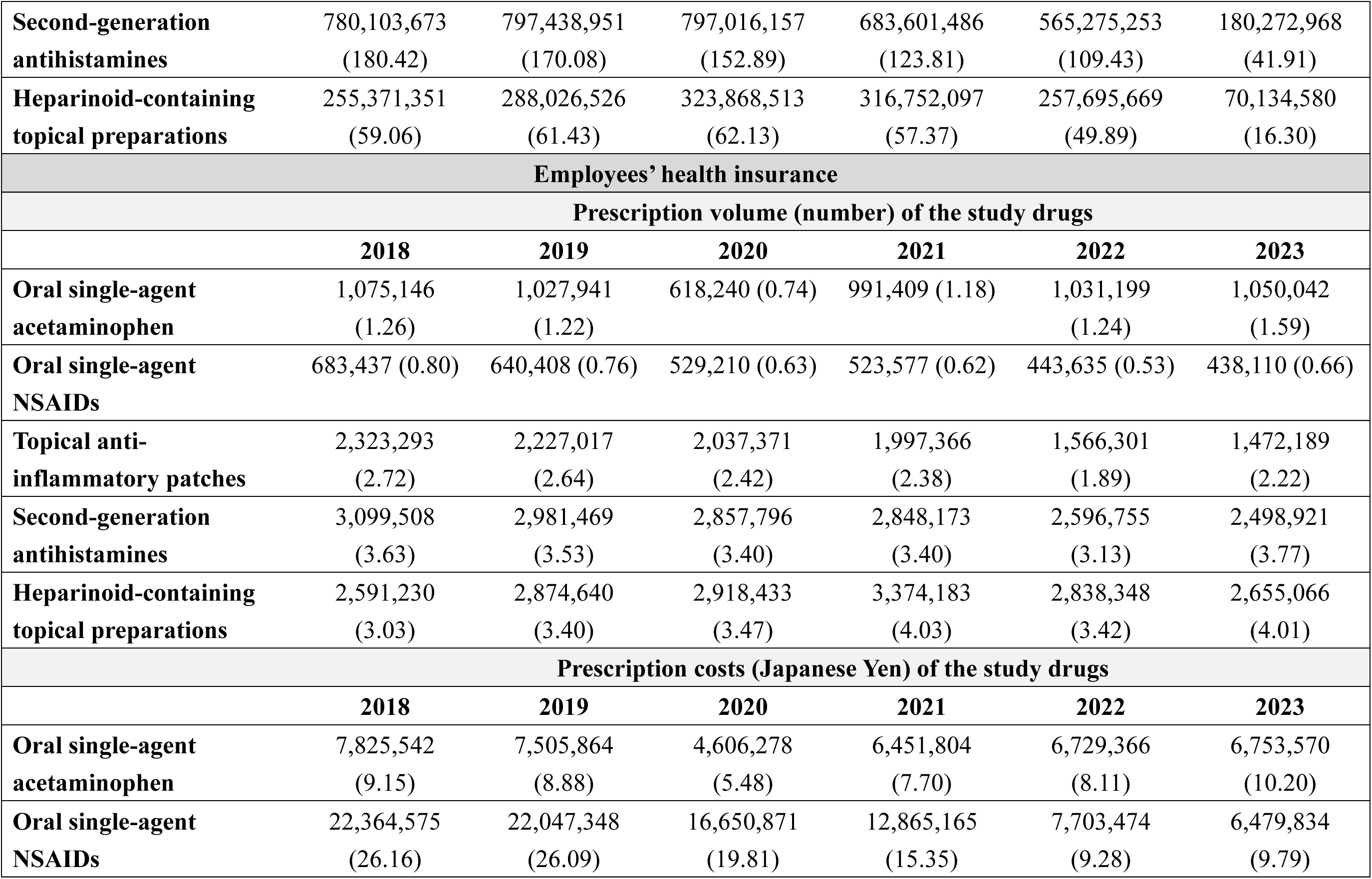
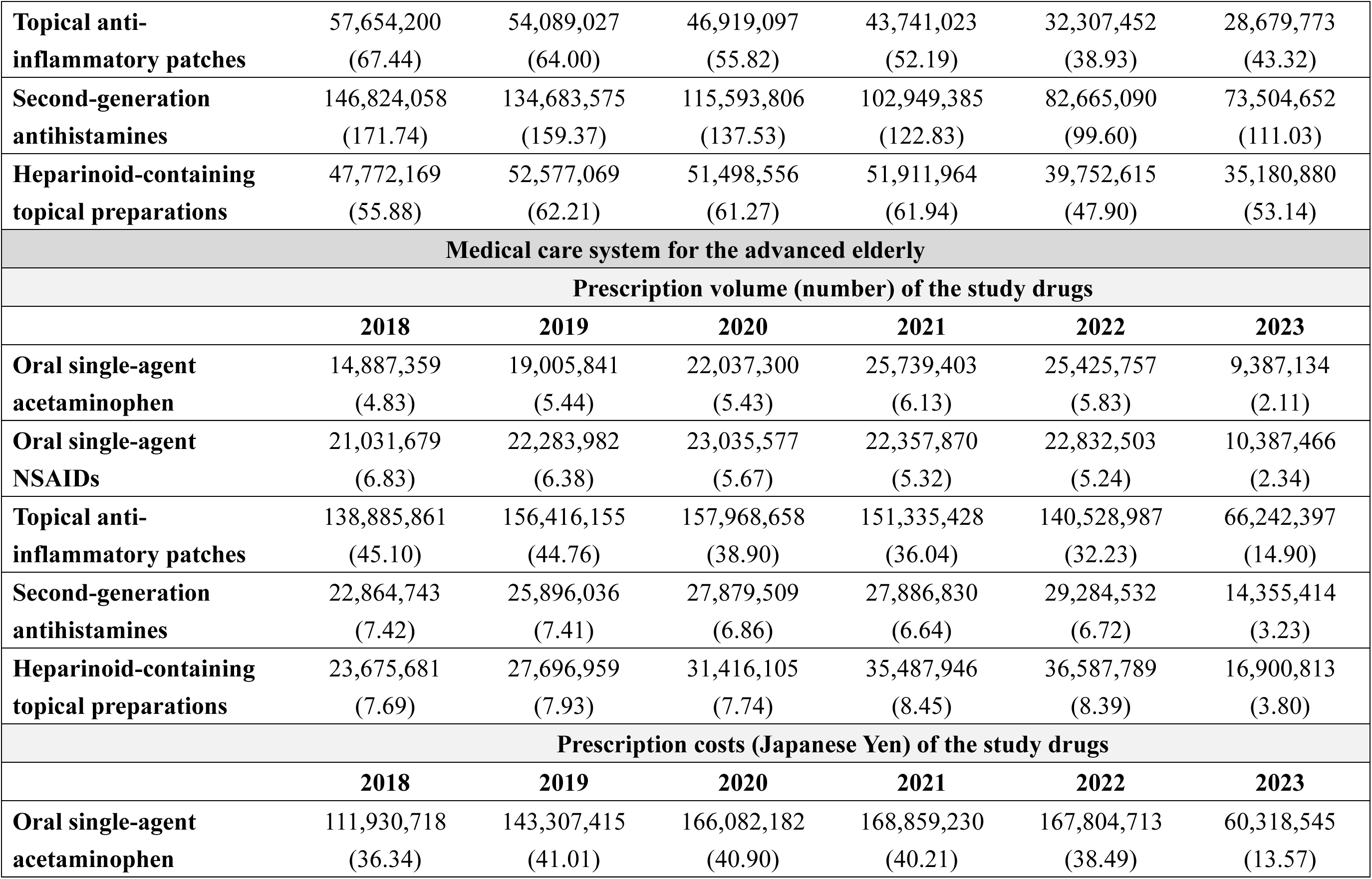
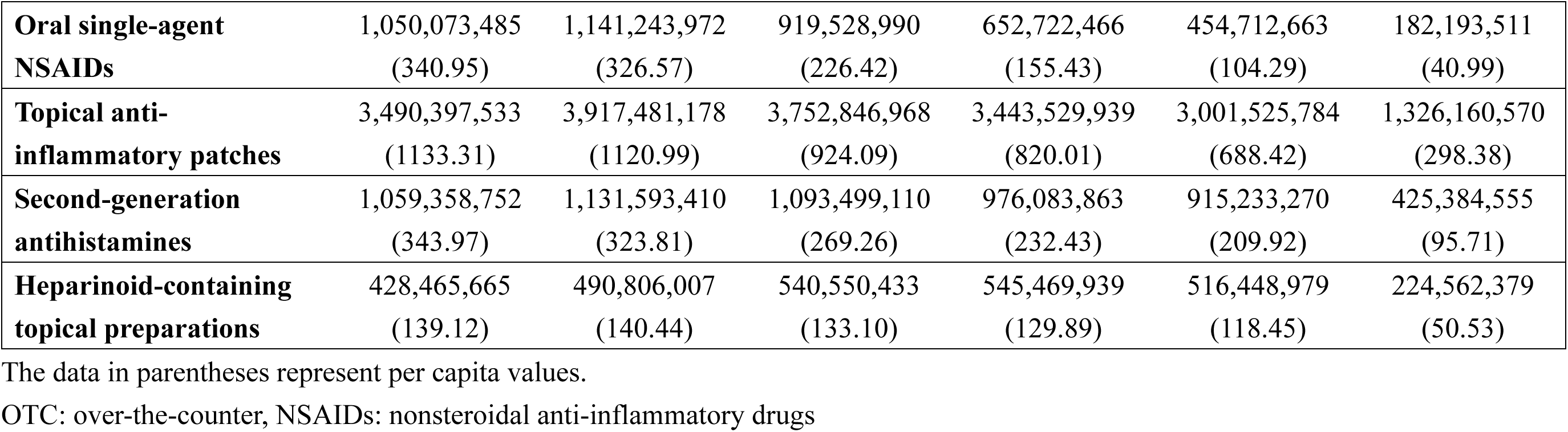
Annual prescription volume and costs of representative OTC-like drugs stratified by health insurance types: Analysis of the DeSC database.

**Supplementary Table 10.** Annual prescription volume and costs of representative OTC-like drugs stratified by healthcare facility types: Analysis of the DeSC database.

| **Clinics** |  |  |  |  |  |  |
| --- | --- | --- | --- | --- | --- | --- |
| **Prescription volume (number) of the study drugs** |  |  |  |  |  |  |
|  | **2018** | **2019** | **2020** | **2021** | **2022** | **2023** |
| **Oral single-agent acetaminophen** | 12,626,587<br>(2.03) | 15,354,487<br>(2.29) | 15,843,486<br>(2.18) | 19,739,659<br>(2.66) | 18,714,059<br>(2.55) | 7,639,996<br>(1.91) |
| **Oral single-agent NSAIDs** | 21,810,891<br>(3.51) | 23,010,689<br>(3.43) | 24,164,039<br>(3.32) | 23,500,508<br>(3.17) | 22,825,822<br>(3.11) | 10,513,274<br>(2.63) |
| **Topical anti-inflammatory patches** | 146,057,406<br>(23.53) | 162,507,481<br>(24.22) | 165,957,948<br>(22.80) | 159,979,614<br>(21.56) | 145,224,643<br>(19.79) | 67,656,235<br>(16.95) |
| **Second-generation antihistamines** | 34,983,348<br>(5.64) | 38,236,839<br>(5.70) | 41,637,102<br>(5.72) | 40,478,513<br>(5.45) | 40,334,754<br>(5.50) | 19,074,037<br>(4.78) |
| **Heparinoid-containing topical preparations** | 28,743,251<br>(4.63) | 33,350,808<br>(4.97) | 37,956,973<br>(5.22) | 42,240,372<br>(5.69) | 40,924,719<br>(5.58) | 18,486,952<br>(4.63) |
| **Prescription costs (Japanese Yen) of the study drugs** |  |  |  |  |  |  |
|  | **2018** | **2019** | **2020** | **2021** | **2022** | **2023** |
| **Oral single-agent acetaminophen** | 93,870,925<br>(15.12) | 114,754,790<br>(17.11) | 118,897,429<br>(16.34) | 128,793,931<br>(17.36) | 122,331,865<br>(16.67) | 48,375,781<br>(12.12) |
| **Oral single-agent NSAIDs** | 957,076,012<br>(154.21) | 1,041,413,882<br>(155.23) | 868,443,041<br>(119.33) | 633,972,672<br>(85.43) | 432,495,966<br>(58.93) | 173,767,458<br>(43.54) |
| **Topical anti-inflammatory patches** | 3,626,635,134<br>(584.34) | 4,019,799,792<br>(599.19) | 3,901,619,584<br>(536.12) | 3,603,958,815<br>(485.64) | 3,076,379,645<br>(419.20) | 1,347,154,643<br>(337.52) |
| **Second-generation antihistamines** | 1,595,078,308<br>(257.01) | 1,648,983,521<br>(245.80) | 1,599,728,329<br>(219.82) | 1,391,582,655<br>(187.52) | 1,238,081,657<br>(168.71) | 549,667,560<br>(137.72) |
| **Heparinoid-containing topical preparations** | 503,245,416<br>(81.09) | 572,180,821<br>(85.29) | 626,112,359<br>(86.03) | 618,961,339<br>(83.41) | 542,753,813<br>(73.96) | 233,138,722<br>(58.41) |
| **Non-university hospitals** |  |  |  |  |  |  |
| **Prescription volume (number) of the study drugs** |  |  |  |  |  |  |
|  | **2018** | **2019** | **2020** | **2021** | **2022** | **2023** |
| **Oral single-agent acetaminophen** | 9,084,944<br>(2.44) | 10,910,488<br>(2.68) | 12,621,901<br>(2.89) | 14,730,688<br>(3.26) | 14,801,787<br>(3.29) | 5,387,308<br>(2.22) |
| **Oral single-agent NSAIDs** | 7,217,492<br>(1.94) | 7,426,613<br>(1.82) | 7,438,035<br>(1.70) | 7,261,750<br>(1.61) | 7,243,677<br>(1.61) | 2,845,452<br>(1.17) |
| **Topical anti-inflammatory patches** | 31,010,589<br>(8.34) | 33,965,440<br>(8.34) | 33,943,529<br>(7.76) | 32,229,397<br>(7.13) | 28,063,813<br>(6.23) | 11,365,706<br>(4.67) |
| **Second-generation antihistamines** | 6,589,660<br>(1.77) | 7,370,685<br>(1.81) | 8,117,225<br>(1.86) | 8,214,881<br>(1.82) | 8,106,843<br>(1.80) | 3,437,115<br>(1.41) |
| **Heparinoid-containing topical preparations** | 8,579,278<br>(2.31) | 10,055,018<br>(2.47) | 11,620,328<br>(2.66) | 12,975,702<br>(2.87) | 12,913,360<br>(2.87) | 5,106,541<br>(2.10) |
| **Prescription costs (Japanese Yen) of the study drugs** |  |  |  |  |  |  |
|  | **2018** | **2019** | **2020** | **2021** | **2022** | **2023** |
| **Oral single-agent acetaminophen** | 67,690,730<br>(18.21) | 81,644,281<br>(20.04) | 94,852,854<br>(21.69) | 96,586,321<br>(21.38) | 98,020,145<br>(21.77) | 35,278,352<br>(14.51) |
| **Oral single-agent NSAIDs** | 415,984,026<br>(111.89) | 436,182,025<br>(107.04) | 343,249,521<br>(78.49) | 224,498,062<br>(49.70) | 148,025,668<br>(32.88) | 55,058,089<br>(22.64) |
| **Topical anti-inflammatory patches** | 806,486,365<br>(216.93) | 871,497,049<br>(213.87) | 817,134,474<br>(186.85) | 745,054,778<br>(164.93) | 609,263,471<br>(135.34) | 236,944,090<br>(97.43) |
| **Second-generation antihistamines** | 321,268,815<br>(86.41) | 342,776,690<br>(84.12) | 341,594,862<br>(78.11) | 304,929,115<br>(67.50) | 266,320,867<br>(59.16) | 110,460,160<br>(45.42) |
| **Heparinoid-containing topical preparations** | 159,772,778<br>(42.98) | 183,505,712<br>(45.03) | 206,814,200<br>(47.29) | 207,892,867<br>(46.02) | 189,694,886<br>(42.14) | 72,784,004<br>(29.93) |
| **University hospitals** |  |  |  |  |  |  |
| **Prescription volume (number) of the study drugs** |  |  |  |  |  |  |
|  | **2018** | **2019** | **2020** | **2021** | **2022** | **2023** |
| **Oral single-agent acetaminophen** | 1,032,826<br>(2.12) | 1,238,627<br>(2.29) | 1,306,413<br>(2.29) | 1,580,090<br>(2.72) | 1,626,076<br>(2.76) | 640,427 (2.04) |
| **Oral single-agent NSAIDs** | 907,844 (1.86) | 867,975 (1.61) | 860,938 (1.51) | 920,161 (1.59) | 928,888 (1.58) | 314,571 (1.00) |
| **Topical anti-inflammatory patches** | 2,488,296<br>(5.11) | 2,555,241<br>(4.73) | 2,440,074<br>(4.28) | 2,542,234<br>(4.38) | 2,283,270<br>(3.87) | 930,306 (2.96) |
| **Second-generation antihistamines** | 1,337,709<br>(2.75) | 1,442,731<br>(2.67) | 1,447,553<br>(2.54) | 1,565,497<br>(2.70) | 1,550,391<br>(2.63) | 541,847 (1.72) |
| **Heparinoid-containing topical preparations** | 3,142,110<br>(6.45) | 3,505,660<br>(6.49) | 3,879,171<br>(6.81) | 4,340,625<br>(7.48) | 4,250,783<br>(7.21) | 1,377,912<br>(4.38) |
| **Prescription costs (Japanese Yen) of the study drugs** |  |  |  |  |  |  |
|  | **2018** | **2019** | **2020** | **2021** | **2022** | **2023** |
| **Oral single-agent acetaminophen** | 8,320,946<br>(17.09) | 10,018,576<br>(18.54) | 10,681,827<br>(18.76) | 11,357,306<br>(19.58) | 11,682,651<br>(19.81) | 4,250,778<br>(13.51) |
| **Oral single-agent NSAIDs** | 52,662,558<br>(108.18) | 52,252,543<br>(96.72) | 39,920,660<br>(70.10) | 32,735,155<br>(56.43) | 20,987,409<br>(35.59) | 5,212,671<br>(16.57) |
| **Topical anti-inflammatory patches** | 75,509,972<br>(155.11) | 76,592,794<br>(141.77) | 69,773,931<br>(122.53) | 68,943,475<br>(118.84) | 54,716,114<br>(92.79) | 21,126,985<br>(67.16) |
| **Second-generation antihistamines** | 67,385,396<br>(138.42) | 70,424,275<br>(130.35) | 63,814,459<br>(112.06) | 65,419,366<br>(112.77) | 58,020,301<br>(98.39) | 18,711,912<br>(59.49) |
| **Heparinoid-containing topical preparations** | 67,338,614<br>(138.33) | 74,901,937<br>(138.64) | 82,469,701<br>(144.82) | 87,073,625<br>(150.09) | 81,288,634<br>(137.85) | 23,920,216<br>(76.04) |

The data in parentheses represent per capita values.
OTC: over-the-counter, NSAIDs: nonsteroidal anti-inflammatory drugs

**Supplementary Figure 1.**
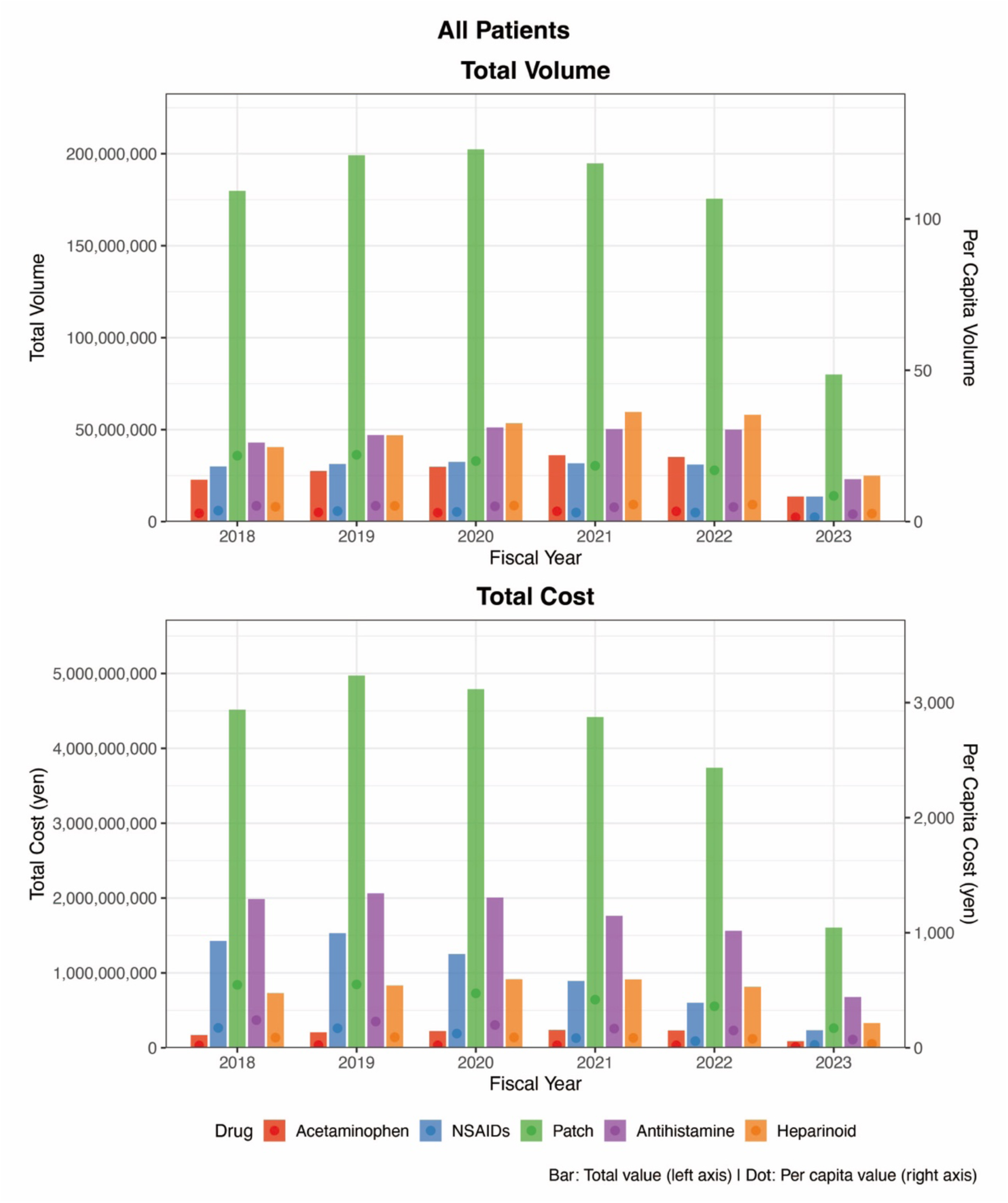
Trends in the prescription volume and costs of representative OTC-like drugs in the overall population: Analysis of the DeSC database. The upper panel shows the total prescription volume (bars, left y-axis) and per capita prescription volume (dots, right y-axis), and the lower panel shows the total prescription costs (bars, left y-axis) and per capita prescription costs (dots, right y-axis). Note: Prescription volume and costs are presented for oral single-agent acetaminophen, oral single-agent nonsteroidal anti-inflammatory drugs (NSAIDs), topical anti-inflammatory patches, oral second-generation antihistamines, and heparinoid-containing topical preparations. OTC: over-the-counter

**Supplementary Figure 2.**
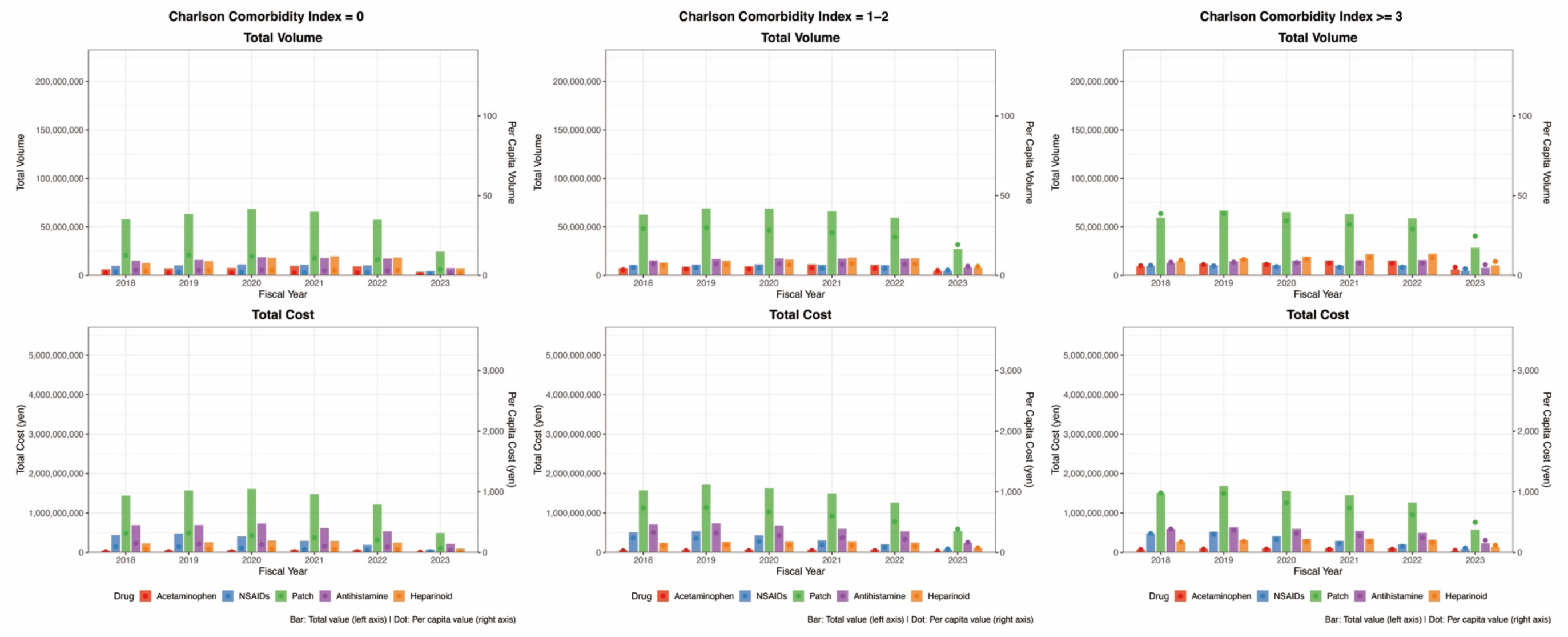
CCI-specific trends in the prescription volume and costs of representative OTC-like drugs: Analysis of the DeSC database. (A) Trends in the total prescription volume (bars, left y-axis) and per capita prescription volume (dots, right y-axis) among individuals with a Charlson Comorbidity Index (CCI) of 0. (B) Trends in the total prescription volume (bars, left y-axis) and per capita prescription volume (dots, right y-axis) among individuals with a CCI of 1–2. (C) Trends in the total prescription volume (bars, left y-axis) and per capita prescription volume (dots, right y-axis) among individuals with a CCI of ≥3. Note: Prescription volume and costs are presented for oral single-agent acetaminophen, oral single-agent nonsteroidal anti-inflammatory drugs (NSAIDs), topical anti-inflammatory patches, oral second-generation antihistamines, and heparinoid-containing topical preparations. OTC: over-the-counter

**Supplementary Figure 3.**
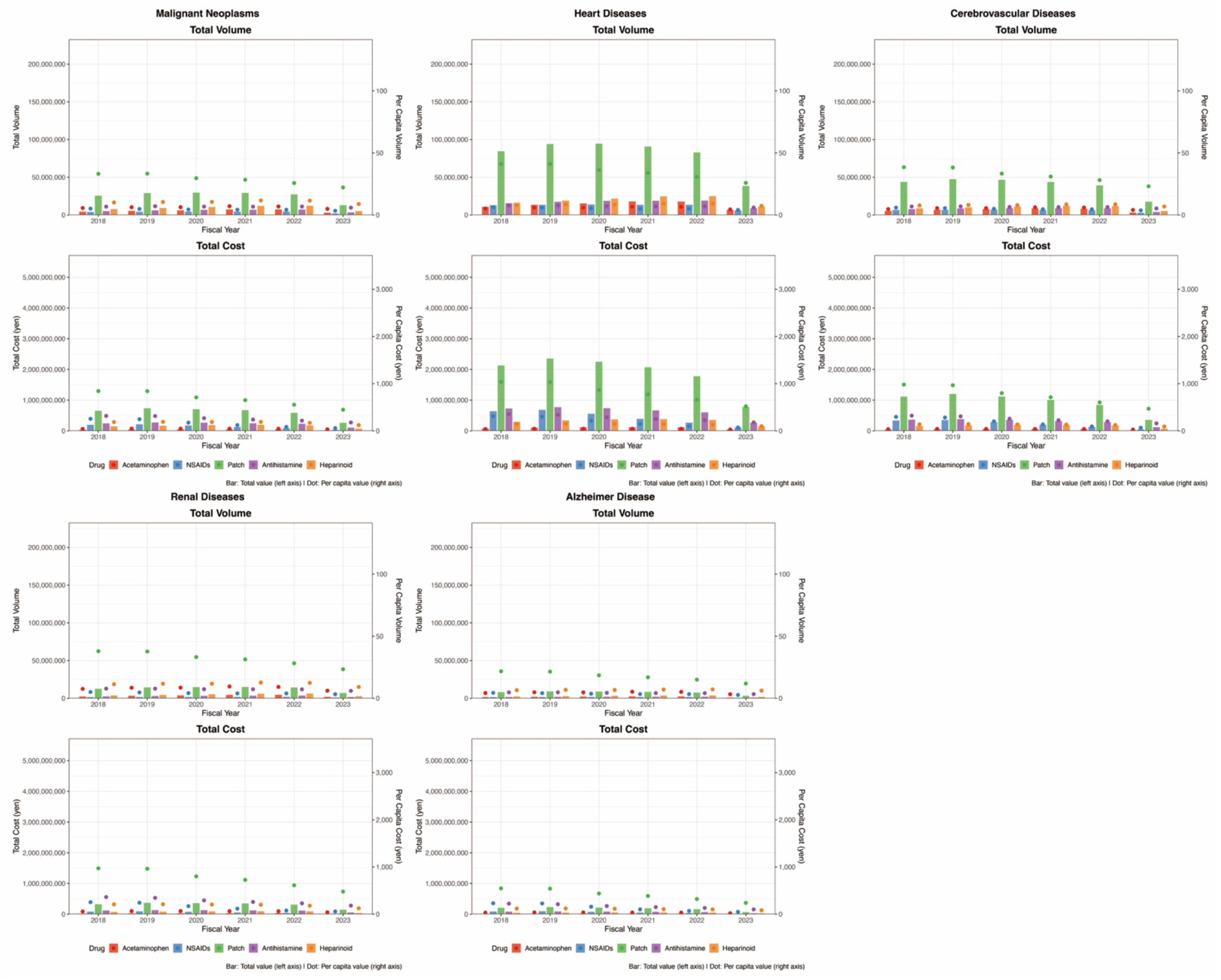
Major-disease–specific trends in the prescription volume and costs of representative OTC-like drugs: Analysis of the DeSC database. (A) Trends in the prescription volume and costs of representative OTC-like drugs in individuals with malignant neoplasms. (B) Trends in the prescription volume and costs of representative OTC-like drugs in individuals with heart diseases. (C) Trends in the prescription volume and costs of representative OTC-like drugs in individuals with cerebrovascular diseases. (D) Trends in the prescription volume and costs of representative OTC-like drugs in individuals with renal diseases. (E) Trends in the prescription volume and costs of representative OTC-like drugs in individuals with Alzheimer’s disease. Note: The bars indicate total values (left y-axis), and dots indicate per capita values (right y-axis). OTC: over-the-counter

**Supplementary Figure 4.**
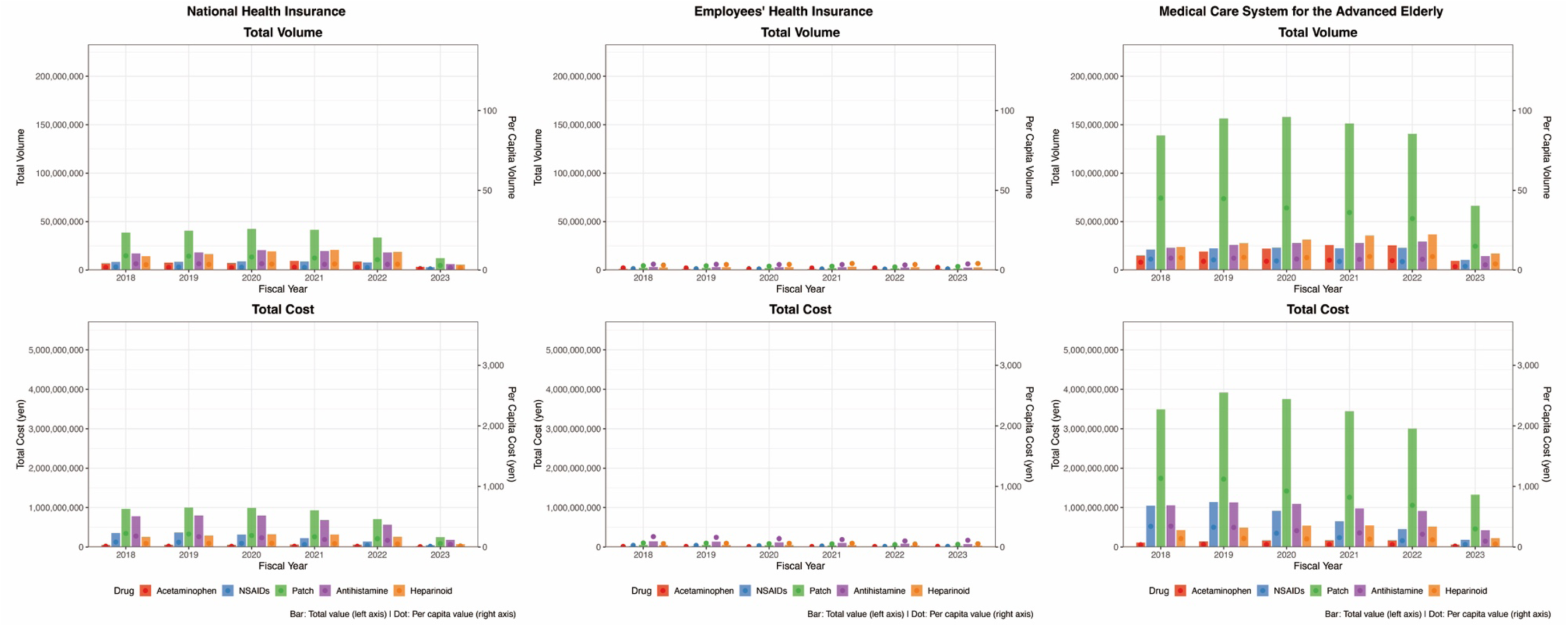
Health-insurance–disease-specific trends in the prescription volume and costs of representative OTC-like drugs: Analysis of the DeSC database. (A) Trends in the prescription volume and costs of representative OTC-like drugs in individuals enrolled in National Health Insurance. (B) Trends in the prescription volume and costs of representative OTC-like drugs in individuals enrolled in Employees’ Health Insurance. (C) Trends in the prescription volume and costs of representative OTC-like drugs in individuals enrolled in the Medical Care System for the Advanced Elderly. Note: The bars indicate total values (left y-axis), and dots indicate per capita values (right y-axis). OTC: over-the-counter

**Supplementary Figure 5.**
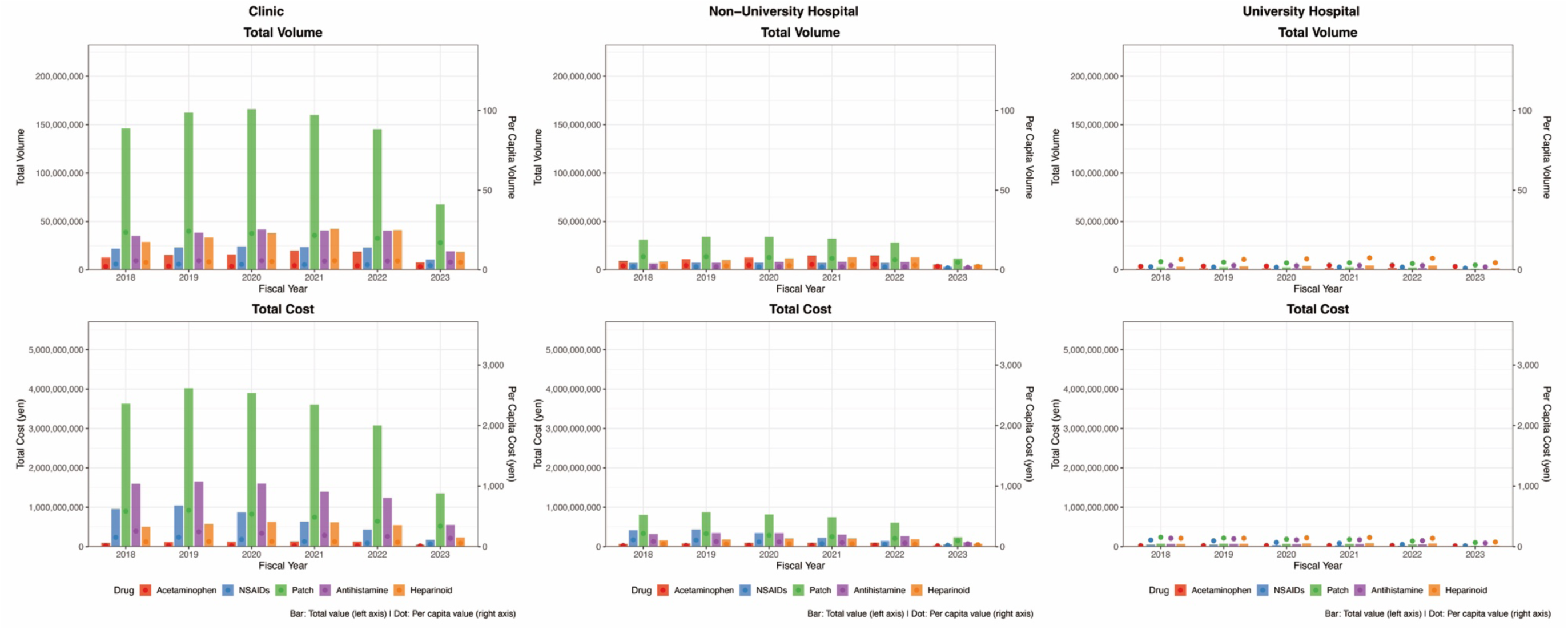
Healthcare-facility–disease-specific trends in the prescription volume and costs of representative OTC-like drugs: Analysis of the DeSC database. (A) Trends in the prescription volume and costs of representative OTC-like drugs in individuals visiting clinics. (B) Trends in the prescription volume and costs of representative OTC-like drugs in individuals visiting non-university hospitals. (C) Trends in the prescription volume and costs of representative OTC-like drugs in individuals visiting university hospitals. Note: The bars indicate total values (left y-axis), and dots indicate per capita values (right y-axis). OTC: over-the-counter

